# Obesity Differs from Diabetes Mellitus in Antibody and T Cell Responses Post COVID-19 Recovery

**DOI:** 10.1101/2023.06.14.23291375

**Authors:** Mohammad Ali, Stephanie Longet, Isabel Neale, Patpong Rongkard, Forhad Uddin Hassan Chowdhury, Jennifer Hill, Anthony Brown, Stephen Laidlaw, Tom Tipton, Ashraful Hoque, Nazia Hassan, Carl-Philipp Hackstein, Sandra Adele, Hossain Delowar Akther, Priyanka Abraham, Shrebash Paul, Md Matiur Rahman, Md Masum Alam, Shamima Parvin, Forhadul Hoque Mollah, Md Mozammel Hoque, Shona C Moore, Subrata K Biswas, Lance Turtle, Thushan I de Silva, Ane Ogbe, John Frater, Eleanor Barnes, Adriana Tomic, Miles W Carroll, Paul Klenerman, Barbara Kronsteiner, Fazle Rabbi Chowdhury, Susanna J Dunachie

## Abstract

**Objective:** Obesity and type 2 diabetes (DM) are risk factors for severe COVID-19 outcomes, which disproportionately affect South Asian populations. This study aims to investigate the humoral and cellular immune responses to SARS-CoV-2 in adult COVID-19 survivors with obesity and DM in Bangladesh.

**Methods:** In this cross-sectional study, SARS-CoV-2-specific antibody and T cell responses were investigated in 63 healthy and 75 PCR-confirmed COVID-19 recovered individuals in Bangladesh, during the pre-vaccination first wave of the COVID-19 pandemic in 2020.

**Results:** In COVID-19 survivors, SARS-CoV-2 infection induced robust antibody and T cell responses, which correlated with disease severity. After adjusting for age, sex, DM status, disease severity, and time since onset of symptoms, obesity was associated with decreased neutralising antibody titers, and increased SARS-CoV-2 spike-specific IFN-γ response along with increased proliferation and IL-2 production by CD8+ T cells. In contrast, DM was not associated with SARS-CoV-2-specific antibody and T cell responses after adjustment for obesity and other confounders.

**Conclusions:** Obesity is associated with lower neutralising antibody levels and higher T cell responses to SARS-CoV-2 post COVID-19 recovery, while antibody or T cell responses remain unaltered in DM.

**Study Importance:** 

**What is already known about this subject?:**

- The impact of obesity and diabetes mellitus (DM) on COVID-19 severity and mortality is disproportionately high in South Asian populations.
- People with obesity and DM experience reduced protection against COVID-19 after vaccination.

**What are the new findings in your manuscript?:**

- Despite similar IgG antibody levels, adults with overweight/obesity (BMI ≥ 23 kg/m^2^) have lower neutralising antibody capacity and higher T cell responses to SARS-CoV-2 following COVID-19 recovery.
- Antigen-specific antibody and T cell responses are preserved in individuals with DM who survive SARS-CoV-2 infection.

**How might your results change the direction of research or the focus of clinical practice?:**

- Our findings underscore the critical need to understand the mechanisms underlying the diminished neutralising capacity of antibodies in obesity, as this has profound implications for the development of effective interventions and treatments for COVID-19.
- Our study highlights the significance of T cells in COVID-19 survivors with obesity, indicating their potential role in informing the development of next-generation vaccines against coronaviruses.

## Introduction

Since the onset of the coronavirus disease 2019 (COVID-19) pandemic, obesity and type 2 diabetes (DM) have emerged as clinically significant risk factors for disease severity, hospitalisation, and mortality due to COVID-19 (1–5). Both comorbidities are associated with increased susceptibility, severity, and poor prognosis in bacterial and other viral infections (6–9). Obesity and DM exhibit similar clinical characteristics including low-grade chronic inflammation (10, 11), impaired energy homeostasis (11, 12), oxidative stress (11, 13), altered cytokine profile (14), and impaired cellular immunity (15, 16), which can significantly impact the body’s ability to fight against pathogens. However, the impact of obesity and DM on COVID-19 remains unclear. Cellular stress, systemic inflammation, and endothelial damage present in metabolic diseases are thought to be aggravated by SARS-CoV-2 infection, increasing the chance of thromboembolism and damage to vital organs (17). Higher expression levels of angiotensin-converting enzyme 2 (ACE2), used for entry of SARS-CoV-2 into cells, are observed in adipose tissue from individuals with obesity and DM (18). Consequently, adipose tissue may act as a reservoir for the virus, promoting a persistent inflammatory response and poor prognosis (19).

Ethnicity has also been linked with COVID-19 severity and mortality. The Office for National Statistics in England reported that COVID-19 mortality in the UK was up to 5 times higher for people of Bangladeshi ethnic background compared to people with White British background, and rate of deaths involving COVID-19 has remained highest for the Bangladeshi ethnic group since the 2^nd^ wave of the pandemic (20, 21). Multiple contributing factors have been proposed to explain worse COVID-19 outcomes in certain ethnicities, with differences related to metabolism and metabolic health amongst these (22) alongside socio-economic factors. Emerging data show ethnicity may modify the association between body mass index (BMI) and COVID-19 outcomes (23, 24). South Asian ethnicities with a BMI of 27 kg/m^2^ have the same risk of COVID-19 mortality as white ethnicities at a BMI of 40 kg/m^2^ (25).

An in-depth understanding of adaptive immune responses to SARS-CoV-2 in South Asian ethnicities, particularly in the Bangladeshi population, is critical to understand the mechanisms associated with disease severity, while they are disproportionately burdened with obesity and DM. This observational clinical study presents our comprehensive analysis of antibody, B cell, and T cell responses to SARS-CoV-2 in COVID-19 survivors in Bangladesh, exploring potential alterations in adaptive immunity related to obesity and DM.

## Methods

### Study design and sample collection

Adult participants aged 18 or over were recruited from Bangabandhu Sheikh Mujib Medical University (BSMMU) Hospital, Dhaka Medical College (DMC) Hospital, and Sheikh Hasina National Institute of Burn and Plastic Surgery (SHNIBPS) in Dhaka, Bangladesh, from September 2020 to November 2020, during Bangladesh’s first wave of COVID-19 pandemic and prior to the global introduction of vaccines. Participants with prior PCR-confirmed SARS-CoV-2 infection with one or more symptoms were recruited at least 28 days after the onset of symptoms. Among them, 31 individuals had recovered from mild/moderate disease (without oxygen support), while 44 individuals had recovered from severe disease (requiring oxygen support). Additionally, 63 healthy control individuals were enrolled who had reported no COVID-19 symptoms since the onset of pandemic. These controls were further categorised as healthy seronegative (n = 35, presumed infection-naive) and healthy seropositive (n = 28, presumed asymptomatic infection) based on anti-spike seropositivity (MSD IgG binding assay – see below).

After obtaining written informed consent, we collected clinical information including age, sex, body mass index (BMI), comorbidities, the date of any SARS-CoV-2 infection (defined by a positive PCR test), presence of symptoms, lowest recorded SpO_2_ during infection (measured by pulse oximeter) and time since onset of symptoms. HbA1c levels were measured by ion-exchange liquid chromatography in a Bio-Rad D-10™ analyser (Bio-Rad Laboratories Inc., Hercules, CA, USA) to assess glycaemic status. Diabetes was defined as HbA1c ≥ 6.5%, while any previous history of diabetes and HbA1c less than 6.5% was included as controlled diabetes. Type 1 and type 2 diabetes were not formally typed, but type 2 diabetes accounts for 90–95% of diabetes cases in Bangladesh (26). Key demographic information is shown in Table 1.

**Table 1.** Demographic characteristics of participants enrolled from September 2020 to November 2020

| **Variable** | **Healthy Seronegative**<br>**(N = 35)** | **Healthy Seropositive**<br>**(N = 28)** | **Recovered (Mild/moderate)**<br>**(N = 31)** | **Recovered (Severe)**<br>**(N = 44)** |
| --- | --- | --- | --- | --- |
| **Age in years, median (IQR)** | 42 (34, 50) | 39 (34, 45) | 33 (28, 42) | 54 (46, 63) |
| **Sex** |  |  |  |  |
| Female, n (%) | 14 (40%) | 9 (32%) | 4 (13%) | 7 (16%) |
| Male, n (%) | 21 (60%) | 19 (68%) | 27 (87%) | 37 (84%) |
| **Diabetes status** |  |  |  |  |
| Non-diabetic, n (%) | 23 (66%) | 13 (46%) | 24 (77%) | 22 (50%) |
| Diabetic, n (%) | 12 (34%) | 15 (54%) | 7 (23%) | 22 (50%) |
| **Obesity category** |  |  |  |  |
| Normal weight, n (%) | 9 (26%) | 6 (21%) | 6 (19%) | 14 (32%) |
| Overweight/obesity, n (%) | 25 (74%) | 21 (76%) | 22 (71%) | 30 (68%) |
| Data missing, n (%) | NA | 1 (3%) | 3 (10%) | NA |
| **Days post symptom onset, median (IQR)** | NA | NA | 43 (32, 122) | 60 (39, 81) |
| IQR (Interquartile range) |  |  |  |  |

The study was approved by the Oxford Tropical Research Ethics Committee (OxTREC reference: 56-20, date: 24/11/2020), the DMC Ethical Review Committee (DMC reference: ERC-DMC/ECC/2020/315, date: 10/09/2020), and the BSMMU Institutional Review Board (BSMMU reference: BSMMU/2020/8760, date: 04/10/2020). For healthy pre-pandemic controls, we used cryopreserved samples (n=40) from a previous unrelated study conducted in Dhaka, Bangladesh in 2017 which was approved by OxTREC (reference: 51-60, date 12/01/2016) and DMC Ethical Review Committee (DMC reference: MEU-DMC/ECC/2015/97(D), date: 04/06/2017). Peripheral blood mononuclear cells (PBMCs) and plasma were separated as previously described (27) and cryopreserved at -80°C then shipped to Oxford, UK on dry ice and stored in liquid nitrogen for later use.

### Meso Scale Discovery (MSD) IgG binding assay

IgG responses to SARS-CoV-2 spike (S), receptor binding domain (RBD) and nucleocapsid (N) antigens were measured using a multiplexed MSD immunoassay: The V-PLEX COVID-19 Coronavirus Panel 3 (IgG) (Meso Scale Discovery, Rockville, MD USA) as previously described (28). Briefly, plasma samples were diluted 1:1,000-30,000 in diluent buffer and added to the MULTI-SPOT® 96-well plates, priorly coated with SARS CoV-2 antigens (S, RBD, N) at 200−400 μg/mL, along with MSD standard and undiluted internal MSD controls. After 2- hour incubation, detection antibody was added. Following washing and addition of read buffer, and plates were read using a MESO® SECTOR S 600 reader. Concentrations are expressed in Arbitrary Units/ml (AU/ml). Cut-offs for each SARS-CoV-2 antigen were as defined previously (27): S, 1160 AU/ml; RBD, 1169 AU/ml; and N, 3874 AU/ml.

### Focus Reduction Neutralisation Test (FRNT)

Plasma was serially diluted in DMEM with 1% FBS from 1:10 to 1:10,000, then combined with equal volume of 100 foci forming units (FFU) of SARS-CoV-2 Victoria virus and incubated for 30 minutes. Vero E6 cells (4.5x10^5/ml) were added 100µl/well and incubated for 2 hours at 37°C, 5% CO2. Carboxymethyl cellulose (1.5%) was then added (100µl/well) and incubated at 37°C, 5% CO2 for 20 hours. Assays were done in duplicate. Cells were washed with DPBS, fixed with 4% paraformaldehyde for 30 minutes at room temperature, and then permeabilised with 1% TritonX100 in PBS, followed by staining with a human monoclonal antibody (FB9B) (29). Bound antibody was detected by incubating with goat anti-human IgG HRP conjugate (Sigma, UK) and followed by TrueBlue™ Peroxidase substrate (Insight Biotechnology, UK), then imaged with an ELISPOT reader. The half-maximal inhibitory concentration (IC50) was defined as the concentration of plasma that reduced the FFU by 50% compared to the control wells.

### Memory B cell Fluorospot assay

Cryopreserved PBMCs were thawed and cultured for 72 hours at 37lll°C, 5% CO2, with polyclonal stimulation containing 1 μg/ml R848 and 10 ng/ml IL-2 from the Human IgA/IgG FluoroSpotFLEX kit (Mabtech) as previously described (30). Stimulated PBMCs were added at 2x10 cells/well to fluorospot plates coated with 10 μg/ml SARS-CoV-2 spike glycoprotein (S) and nucleocapsid protein (both from The Native Antigen Company, UK) diluted in PBS (Gibco). Plates were incubated for 18 hours in a humidified incubator at 37lll°C, 5% CO2, and developed according to the manufacturer’s instructions (Mabtech). Analysis was carried out with AID ELISpot software 8.0 (Autoimmun Diagnostika). Memory B cell IgG response was measured as antibody spot-forming units (SFU) per million.

### T cell interferon-gamma ELISpot Assay

The Standard Operating Procedure for T cell interferon-gamma (IFN-γ) ELISpot Assay has been published previously (28). In brief, cryopreserved PBMCs were thawed and plated at 200,000 cells/well in a MultiScreen-IP filter plate (Millipore, MAIPS4510) previously coated with capture antibody (clone 1-D1K). PBMCs were then incubated with overlapping SARS-CoV-2 peptide pools (18-mers with 10 amino acid overlap, Mimotopes), representing the S1, S2, membrane (M), nucleocapsid (N), open reading frame (ORF) 3 and 6, ORF 7 and 8 (2 ug/ml) for 16 to 18 hours in a humidified incubator at 37lll°C, 5% CO_2_. Positive controls included the cytomegalovirus pp65 protein (Miltenyi Biotec) and concanavalin A, DMSO served as the negative control. After incubation, the plates were developed following manufacturers protocol (Mabtech 3420-2A) and analysed with the ImmunoSpot® S6 Alfa Analyser (Cellular Technology Limited LLC, Germany). Antigen-specific responses were quantified as IFN-γ spot-forming units (SFU)/million PBMC. A positive response was defined as greater than 37 SFU/million PBMC based on the mean + 2 standard deviations (SD) of the negative control wells.

### Proliferation assay

T cell proliferation assay was carried out as previously described (27). In brief, cryopreserved PBMCs were thawed and stained with CellTrace® Violet (Life Technologies). PBMCs were then plated in 96-well plates and stimulated with SARS-CoV-2 peptide pools (S1, S2, M, N, ORF3 and 6, ORF 7 and 8) at 1lllμg/ml. DMSO and PHA-L were used as negative and positive controls, respectively. Cells were incubated at 37lll°C, 5% CO2 for 7 days with media change on day 4. Flow cytometry was used to analyse the relative frequency of proliferating CD4+ and CD8+ T cells. Responses above 1% were considered true positive.

### Intracellular cytokine stimulation assay

In a subset of donors (n=36), selected from healthy seronegative (n=5), healthy seropositive (n=13), and recovered patients (n=18), T cell responses were characterised by intracellular cytokine staining (ICS) as described previously (30). In brief, PBMCs were stimulated with SARS-CoV-2 peptide pools (spike, M, and N) at 2 μg/ml along with anti-CD28 and anti-CD49d (both from BD). DMSO and a cell activation cocktail (PMA and ionomycin) were used as negative and positive controls, respectively. After 1 hour incubation, Brefeldin A was added and the samples were further incubated for 15 hours at 37°C and 5% CO_2_. Flow cytometry was used to analyse the relative frequency of IFN-γ, TNF, and IL-2 producing CD4+ and CD8+ T cells. Details of antibodies used for proliferation assay and ICS are listed in Table S3.

### Statistical analysis

Statistical analyses were conducted using R version 4.2.1 (https://www.R-project.org/). Unpaired comparisons between two or more groups were assessed using the two-tailed Wilcoxon rank-sum test or Kruskal-Wallis test with Dunn’s multiple comparisons test as appropriate. Correlation analysis was performed using Spearman’s rank correlation coefficient. SIMON software version 0.2.1 (https://genular.org) (31) was used to perform correlation and principal component analysis. A significance level of Plll<lll0.05 was set.

The detailed descriptions of Generalised Linear Models (GLMs) are provided in Supporting Information Methods, and summary tables are reported in Table S2a-f. Cases with missing data were eliminated in GLMs.

## Results

### SARS-CoV-2 infection induces robust antibody responses, which are more pronounced in recovery from severe illness

We first analysed SARS-CoV-2-specific IgG responses among our study cohorts (Figure 1A) using the MSD IgG binding assay. Participants who recovered from severe disease had significantly higher IgG responses to SARS-CoV-2 spike, RBD, and N than those who recovered from mild/moderate disease and healthy seropositive controls, with the latter two groups not showing any differences (Figure 1B). Despite these differences, using an ex vivo memory B cell ELISpot assay, we found comparable numbers of memory B cells specific to spike and N in healthy seropositive individuals and those who recovered from mild/moderate and severe disease, while healthy seronegative controls lacked detectable IgG+ memory B cells (Figure 1C) in blood. Further, individuals who recovered from mild/moderate disease tended to have elevated levels of antibody (Ab) neutralisation of SARS-CoV-2 compared to those who recovered from severe disease; however, the difference was not statistically significant (P=0.063) (Figure 1D).

**Figure 1:**
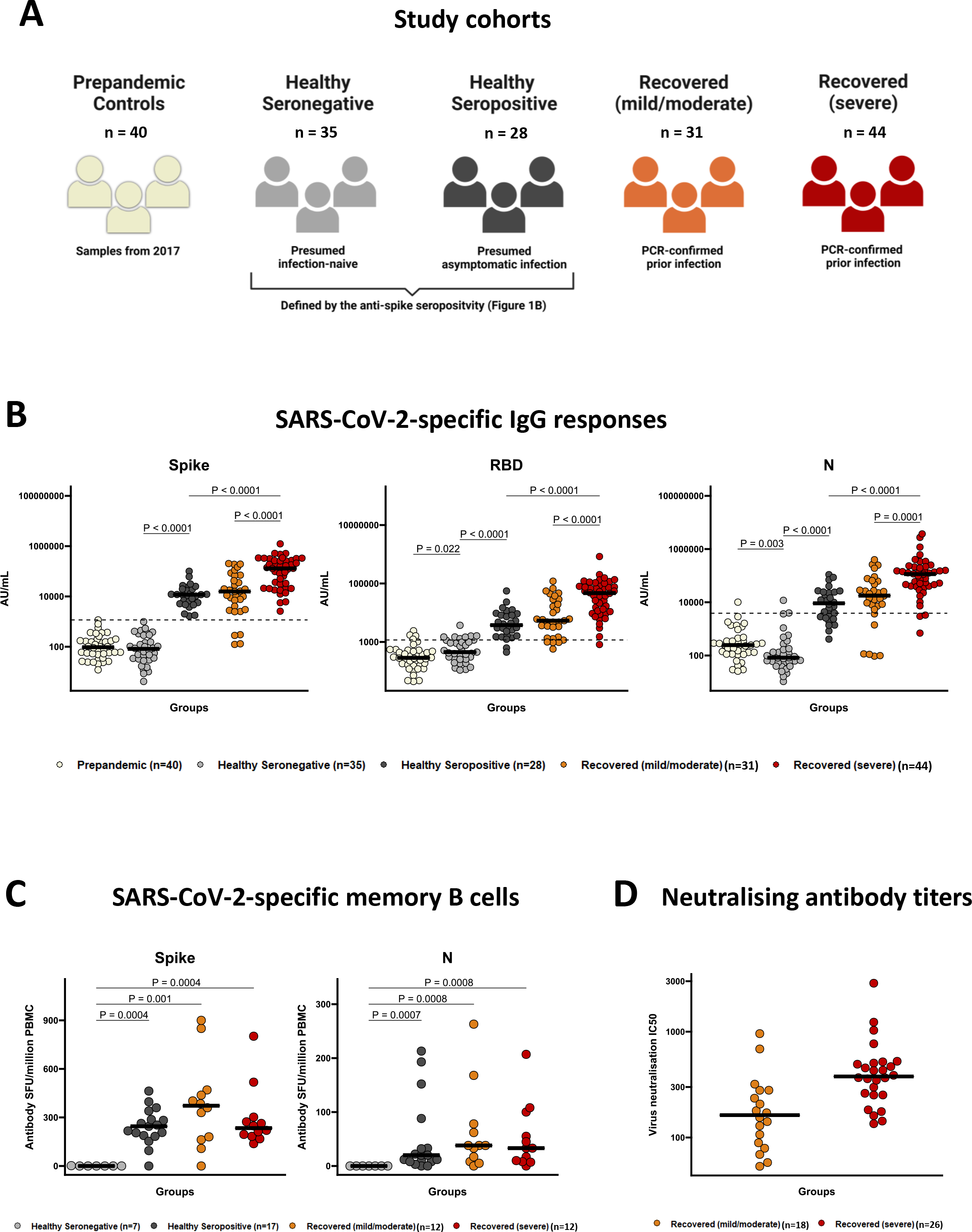
SARS-CoV-2 infection induces robust antibody responses, which are more pronounced in recovery from severe illness. (A) Study cohorts including pre-pandemic controls (n=40; recruited in Bangladesh in 2017 – ivory colour), healthy seronegative controls (n=35, presumably infection naïve - grey colour), healthy seropositive controls (n=28, presumably asymptomatic infection – black colour), individuals who recovered from mild/moderate disease (n=31 – orange colour) and severe disease (n=44 – red colour) due to PCR-confirmed symptomatic SARS-CoV-2 infection and recruited at least 28 days after onset of symptoms. Participants (except pre-pandemic controls) were recruited from September 2020 to November 2020, during Bangladesh’s first wave of the COVID-19 pandemic and prior to the global introduction of vaccines. Seropositivity status was defined by MSD IgG binding assay. (B) IgG responses to SARS-CoV-2 spike (S), receptor binding domain (RBD) and nucleocapsid (N) antigens in pre-pandemic controls (n=40), healthy seronegative controls (n=35), healthy seropositive controls (n=28), individuals recovered from mild/moderate disease (n=31) and severe illness (n=44). IgG responses were measured in plasma samples using multiplexed MSD immunoassays and are expressed in arbitrary units (AU)/mL Horizontal dotted lines represent the cut-off of each assay based on the pre-pandemic sera from UK individuals. (C) Spike and N -specific memory B cell IgG responses in healthy seronegative controls (n=7), healthy seropositive controls (n=17), and individuals recovered from mild/moderate disease (n=12) and severe illness (n=12). Memory B cells were quantified by B cell ELISpot assay from cryopreserved peripheral blood mononuclear cells (PBMC), and data are shown in antibody spot-forming units (SFU)/million PBMC. (D) Neutralising antibody titers against SARS-CoV-2 by focus reduction neutralisation (FRNT) assay in individuals recovered from mild/moderate disease (n=18) and severe illness (n=26). IC_50_ is the reciprocal dilution of the concentration of plasma required to produce a 50% reduction in infectious focus-forming units of virus in Vero cells (ATCC, CCL-81). Bars for (B) and (C) represent the medians. Groups were compared with Kruskal-Wallis nonparametric test, with only significant 2-tailed p values (p<0.05) shown above linking lines.

SARS-CoV-2 infection elicits robust T cell responses that correlate with disease severity We assessed T cell responses to SARS-CoV-2 in our study cohorts using an ex vivo IFN-γ ELISpot assay, a proliferation assay, and an intracellular cytokine assay (ICS). Both healthy seropositive individuals and recovered cohorts (mild/moderate and severe disease) exhibited detectable IFN-γ ELISpot responses to SARS-CoV-2 peptide pools spanning structural (S1, S2, M, N) and accessory (ORF3 and 6, ORF7 and 8) proteins, whereas the pre-pandemic controls and most healthy seronegative controls had no detectable responses above the defined positivity threshold (37 SFU/10 PBMC) (Figure 2A, Figure S1A). Subjects who recovered from severe illness mounted substantially greater IFN-γ responses to spike (summed S1 and S2 responses), and ORFs (summed ORF3 and 6, ORF7 and 8) than those who recovered from mild/moderate disease and healthy seropositive controls (Figure 2B). Furthermore, both recovered cohorts had a greater magnitude of IFN-γ responses to M+N compared to the healthy seropositive controls (Figure 2B). Notably, the breadth of IFN-γ responses to SARS-CoV-2 peptide pools was greater in both groups of recovered individuals as compared to healthy seropositive controls (Figure S1A). These results highlight the robustness of anti-SARS-CoV-2 IFN-γ ELISpot responses across different disease severities.

**Figure 2:**
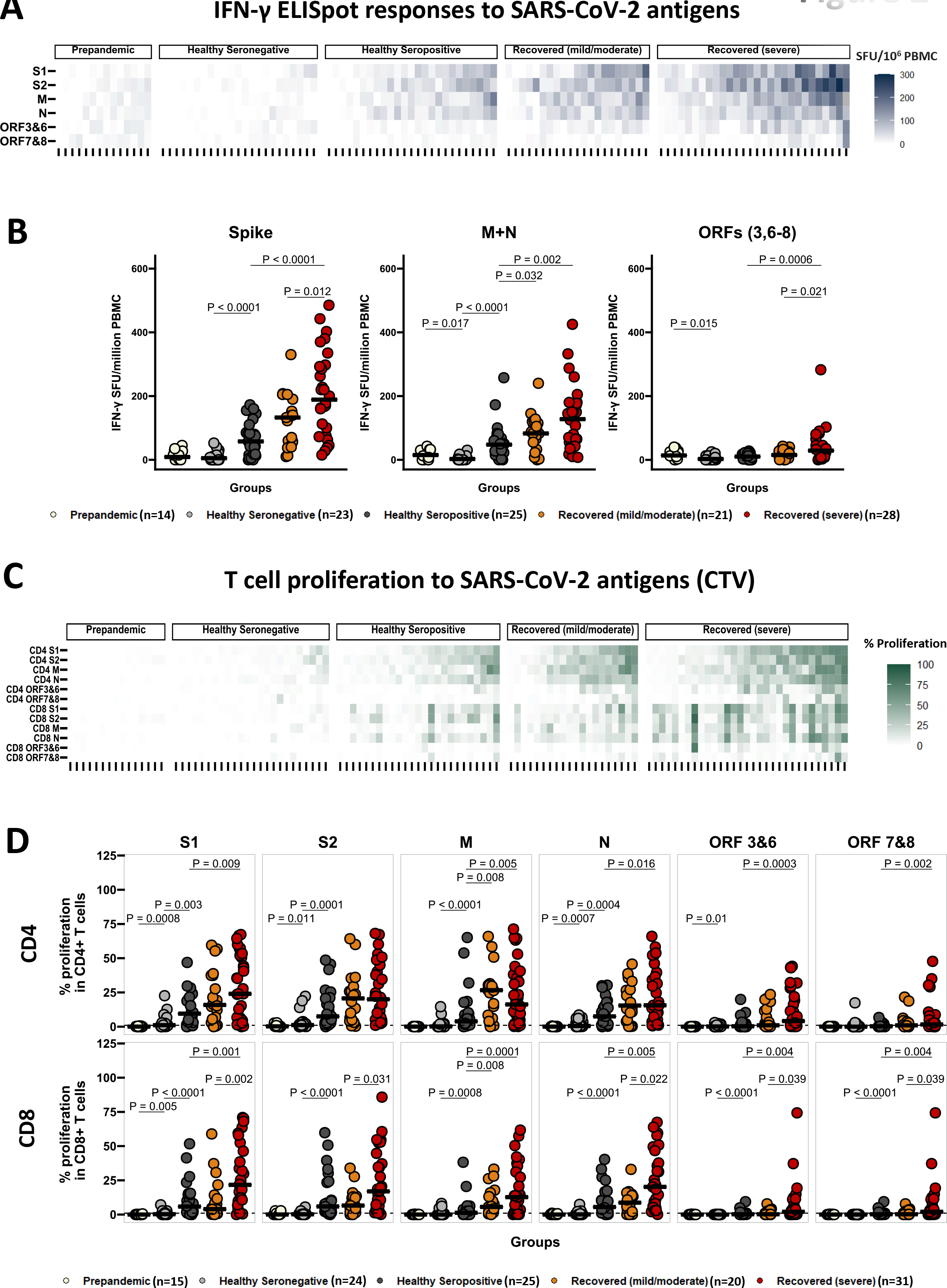
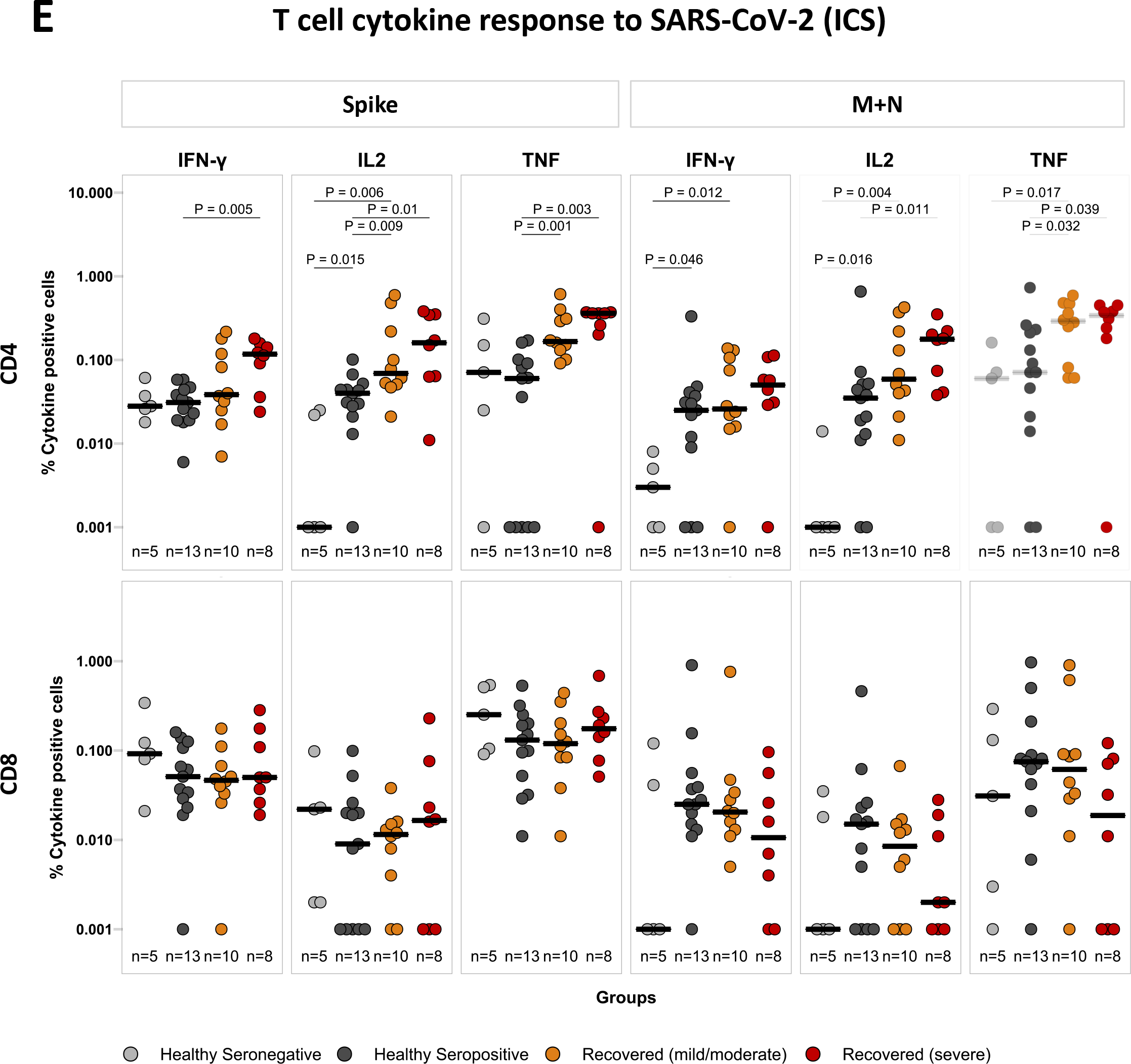
SARS-CoV-2 infection elicits robust T cell responses that correlate with disease severity. (A) Heatmap displaying the IFN-γ responses to SARS-CoV-2 peptide pools spanning structural and accessory proteins (S1, S2, M, N, ORF3 and 6, ORF7 and 8) in pre-pandemic controls (n=14), healthy seronegative controls (n=23), healthy seropositive controls (n=25), individuals recovered from mild/moderate disease (n=21) and severe illness (n=28). IFN-γ responses were measured by ex vivo IFN-γ ELISpot assay from cryopreserved PBMC samples, and data are shown in IFN-γ spot-forming units (SFU)/million PBMC. (B) Comparison of IFN-γ ELISpot responses to SARS-CoV-2 spike (summed responses to S1 and S2 peptide pools), M+N (summed responses to M and N pools), and ORFs (summed responses to ORF3, 6-8) in prepandemic controls (n=14), healthy seronegative controls (n=23), healthy seropositive controls (n=25), individuals recovered from mild/moderate disease (n=21) and severe illness (n=28). (C) Heatmap displaying the relative frequency of CD4^+^ and CD8^+^ T cells proliferating to individual peptide pools S1, S2, M, N, ORF3&6, ORF7&8, assessed by flow cytometry (gating strategy shown in figure S2A) from cryopreserved PBMC, in prepandemic controls (n=15), healthy seronegative controls (n=24), healthy seropositive controls (n=25), individuals recovered from mild/moderate disease (n=20) and severe illness (n=31). (D) Comparison of relative frequency of CD4^+^ (top panels) and CD8^+^ (bottom panels) T cells proliferating to SARS-CoV-2 individual peptide pools in prepandemic controls (n=15), healthy seronegative controls (n=24), healthy seropositive controls (n=25), individuals recovered from mild/moderate disease (n=20) and severe illness (n=31). (E) The spike and M+N -specific IFNγ, IL2, and TNF expression levels are reported as a percentage of the CD4^+^ T cell population (top panels) and CD8^+^ T cell population (bottom panels). Cryopreserved PBMCs from healthy seronegative controls (n=5), healthy seropositive controls (n=13), and individuals recovered from mild/moderate disease (n=10) and severe illness (n=8) were analysed by intracellular cytokine staining and flow cytometry (gating strategy is shown in figure S2B). Bars for (B), (D), and (E) represent the medians. Groups were compared with Kruskal-Wallis nonparametric test, with only significant 2-tailed p values (p<0.05) shown above linking lines.

To obtain a deeper understanding of the induction of T cell based immunity in our cohorts, we next performed proliferation assays, a sensitive method to assess the SARS-CoV-2 specific circulating CD4+ and CD8+ T cells (27). Recovered individuals with severe disease had greater magnitude and breadth of CD4+ and CD8+ T cell proliferative responses to SARS-CoV-2 peptide pools than healthy seropositive controls (Figure 2C-D, Figure S1B-C). Notably, CD8+ T cell proliferation was significantly higher and broader in recovered individuals with severe illness than those who recovered from mild/moderate disease (Figure 2D, Figure S1C). Moreover, healthy seropositive controls displayed a higher magnitude and breadth of T cell proliferative responses in comparison to healthy seronegative controls (Figure 2C-D, Figure S1B-C). Interestingly, there was no detectable T cell proliferation in our pre-pandemic controls (Figure 2C-D, Figure S1B-C), in contrast to UK pre-pandemic controls (27).

In ICS, we observed higher CD4+ T cell IFN-γ, IL-2, and TNF responses to SARS-CoV-2 spike and higher CD4+ T cell IL-2, and TNF responses to M+N in participants who recovered from severe illness compared to the healthy seropositive controls (Figure 2E). Similarly, those who recovered from mild/moderate illness had higher CD4+ T cell IL-2 and TNF responses to spike, and TNF responses to M+N than healthy seropositive controls (Figure 2E). In contrast, we did not detect any differences among the groups in the levels of CD8+ T cell cytokine expressions (Figure 2E).

Individuals with overweight/obesity had lower neutralising antibody titers, but higher T responses to SARS-CoV-2 following recovery We next went on to assess immune features associated with obesity in individuals who had recovered from PCR-confirmed SARS-CoV-2 symptomatic infection. We combined the participants from our two recovered cohorts (mild/moderate and severe disease) and grouped them as lean (BMI = 18.5–22.9 kg/m^2^ ), and overweight/obesity (Ov/Ob, BMI ≥ 23 kg/m^2^ ) following the WHO recommendation for Asian BMI (32). Notably, participants with Ov/Ob were considerably younger than those who were lean (Table S1a).

The recovered participants with Ov/Ob had 2-fold lower neutralising antibody titers compared to the lean subjects, with no differences in the IgG responses to SARS-CoV-2 spike, RBD and N (Figure 3A). In addition, the memory B cell responses to SARS-CoV-2 N were 2-fold lower in Ov/Ob compared to lean (Figure 3A). In contrast, IFN-γ ELISpot responses to SARS-CoV-2 M+N peptide pools were 2-fold higher in the participants with Ov/Ob, with a similar trend (P = 0.0685) in the spike–specific IFN-γ responses (Figure 3B). In ICS, IFN-γ expression by CD4+ T cells was 3-fold higher in response to M+N, and IL-2 expression by CD8+ T cells was 16-fold higher in response to spike in Ov/Ob compared to lean (Figure S3). CD8+ T cell proliferative responses to S1, S2, and N peptide pools were 2 to 4-fold higher in Ov/Ob, while CD4+ T cell proliferation was comparable between lean and Ov/Ob (Figure 3C).

**Figure 3:**
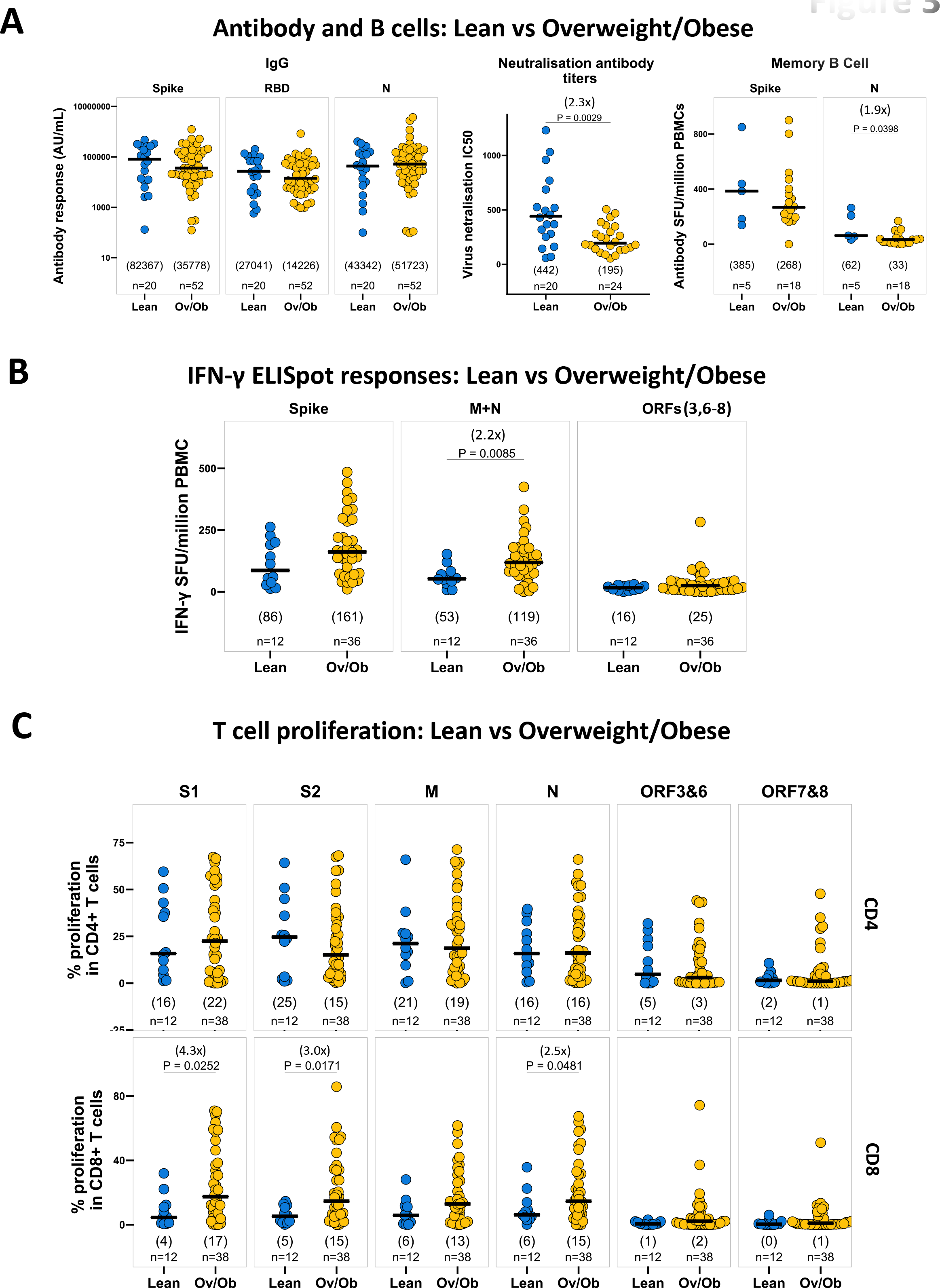
Overweight/obese status is associated with lower neutralising antibody titers and memory B cell responses, but higher T responses to SARS- CoV-2 in patients who recovered from symptomatic SARS-CoV-2 infection. (A) Comparison of SARS-CoV-2 spike, RBD, N -specific IgG responses, neutralising antibody titers, and spike, N -specific memory B cells in lean (BMI = 18.5 – 22.9 kg/m^2^) and overweight/obese (BMI ≥ 23 kg/m^2^) individuals, who recovered from symptomatic SARS-CoV-2 infection. IgG responses, neutralising antibody titers, and memory B cell responses are measured by multiplexed MSD immunoassays, Focus reduction neutralisation (FRNT) assay, and B cell ELISpot assay, respectively, and data are shown in arbitrary units (AU)/mL, IC50, and antibody spot-forming units (SFU)/million PBMC respectively. (B) Comparison of IFN-γ ELISpot responses to SARS-CoV-2 spike (summed responses to S1 and S2 peptide pools), M+N (summed responses to M and N pools), and ORFs (summed responses to ORF3, 6-8) from cryopreserved PBMCs in lean (BMI = 18.5 – 22.9 kg/m^2^) and overweight/obese (BMI ≥ 23 kg/m^2^) individuals in recovery following symptomatic SARS-CoV-2 infection. Data are shown in IFN-γ spot- forming units (SFU)/million PBMC. (C) Comparison of the relative frequency of CD4^+^ (top panels) and CD8^+^ (bottom panels) T cells proliferating to individual peptide pools S1, S2, M, N, ORF3&6, ORF7&8, assessed by flow cytometry (gating strategy shown in figure S2A) from cryopreserved PBMC, in lean (BMI = 18.5 – 22.9 kg/m^2^) and overweight/obese (BMI ≥ 23 kg/m^2^) SARS-CoV-2 recovered patients. A two-tailed Wilcoxon rank-sum test was used to compare between the groups (without correction for multiple testing), and fold changes in brackets referring to the p value comparisons directly below are shown on the top of the dot plots, in case of significant differences. The number of individuals (n) evaluated per assay is displayed at the bottom of the corresponding dot plots. Horizontal bars represent the medians, and the median values are shown in brackets immediately above the number of individuals (n) in each column.

Individuals with diabetes had higher IgG and CD4+ T cell proliferative responses to SARS-CoV-2 following recovery We then compared the immune responses between the recovered individuals with diabetes (DM, HbA1c ≥ 6.5%) and without diabetes (non-DM, HbA1c < 6.5%). It is noteworthy that the individuals with DM were considerably older and had more frequently recovered from severe disease compared to the non-DM individuals (Table S1b).

IgG responses to SARS-CoV-2 spike, RBD, and N were 3 to 8-fold higher in the individuals with DM compared to non-DM (Figure 4A). We also noticed a trend of higher neutralising antibodies (P = 0.082) in the individuals with DM, in contrast to the lower levels seen in obesity, but no difference was observed in either memory B cell responses to SARS-CoV-2 (Figure 4A) or SARS-CoV-2-specific IFN-γ ELISpot responses between individuals with DM and non-DM (Figure 4B). Interestingly, CD4+ T cell proliferation was around 3-fold higher to S1 and ORF 7+8 in individuals with DM, while the proportion of proliferating CD8+ T cells was comparable between the DM and non-DM cohorts (Figure 4C). We did not find any differences in CD4+ and CD8+ cytokine responses by ICS in the DM and non-DM participants.

**Figure 4:**
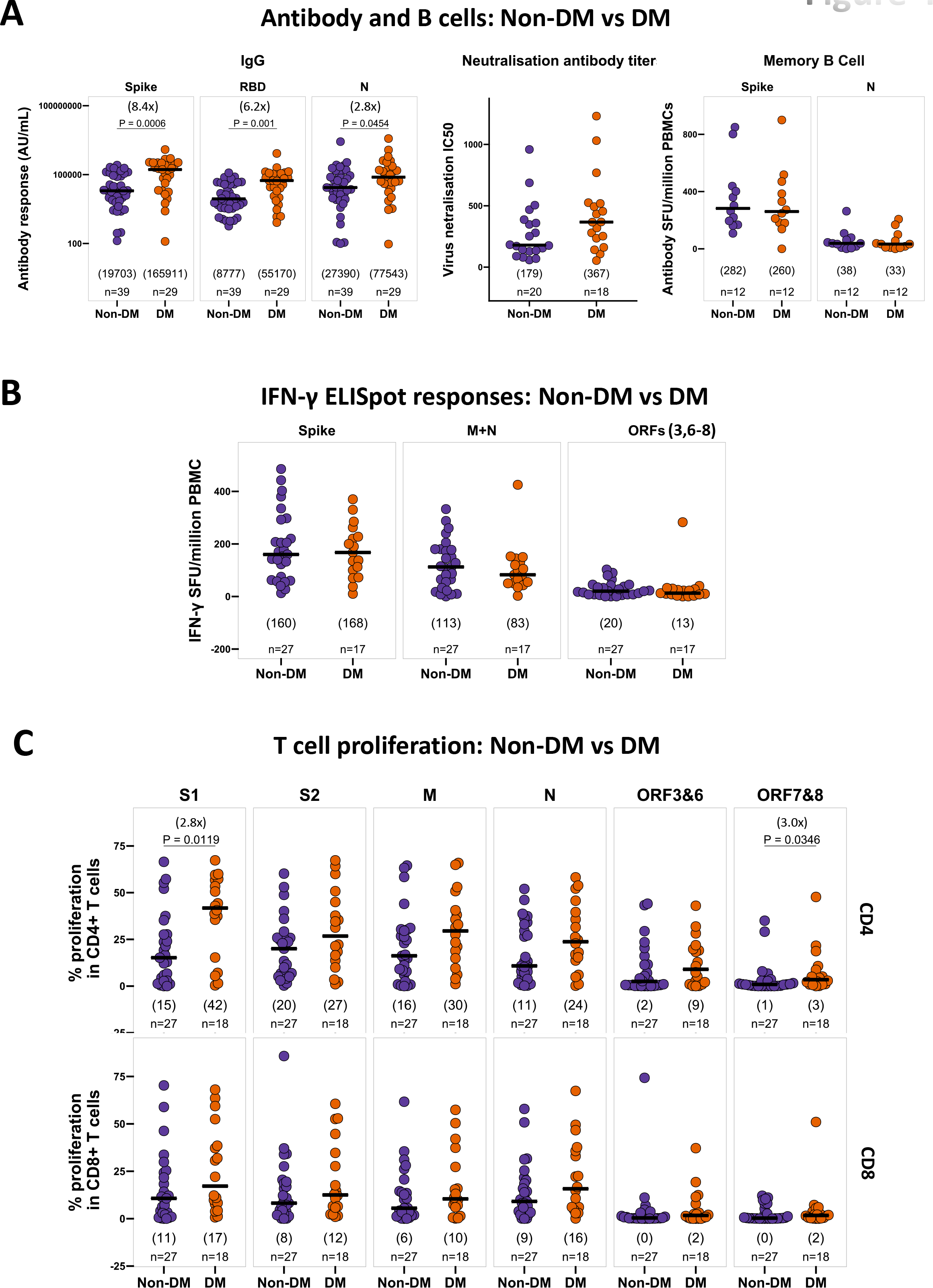
Diabetes is associated with higher IgG responses and CD4+ T cell proliferation to SARS-CoV-2 in patients following recovery from symptomatic SARS-CoV-2 infection. (A) Comparison of SARS-CoV-2 spike, RBD, N -specific IgG responses, neutralising antibody titers, and spike, N -specific memory B cells in non-diabetic (non-DM, HbA1c < 6.5%) and diabetic (DM, HbA1c ≥ 6.5%) individuals, who recovered from symptomatic SARS-CoV-2 infection. IgG responses, neutralising antibody titers, and memory B cell responses are measured by multiplexed MSD immunoassays, focus reduction neutralisation (FRNT) assay, and B cell ELISpot assay, respectively, and data are shown in arbitrary units (AU)/mL, IC_50_, and antibody spot-forming units (SFU)/million PBMC respectively. (B) Comparison of IFN-γ ELISpot responses to SARS-CoV-2 spike (summed responses to S1 and S2 peptide pools), M+N (summed responses to M and N pools), and ORFs (summed responses to ORF3, 6-8) from cryopreserved PBMCs in recovered patients with or without diabetes, following symptomatic SARS-CoV-2 infection. Data are shown in IFN-γ spot-forming units (SFU)/million PBMC. (C) Comparison of the relative frequency of CD4^+^ (top panels) and CD8^+^ (bottom panels) T cells proliferating to individual peptide pools S1, S2, M, N, ORF3&6, ORF7&8, assessed by flow cytometry (gating strategy shown in figure S2A) from cryopreserved PBMC, in SARS-CoV-2 recovered patients with or without diabetes. A two-tailed Wilcoxon rank-sum test was used to compare between the groups (without correction for multiple testing), and fold changes in brackets referring to the p value comparisons directly below are shown on the top of the dot plots, in case of significant differences. The number of individuals (n) evaluated per assay is displayed at the bottom of the corresponding dot plots. Horizontal bars represent the medians, and the median values are shown in brackets immediately above the number of individuals (n) in each column.

Clinical factors were associated with antibody and T cell responses to SARS-CoV-2 We performed a univariable correlation analysis to unravel the impact of a range of variables including age, body mass index (BMI), glycemic status (HbA1c), SpO_2_ during infection, and the time since onset of symptoms (Figure 5A) on the antiviral immune response. Our results show that age was positively correlated with IgG and neutralising antibody titers, IFN-γ ELISpot responses, T cell proliferation, and CD4+ T cell cytokines.

**Figure 5:**
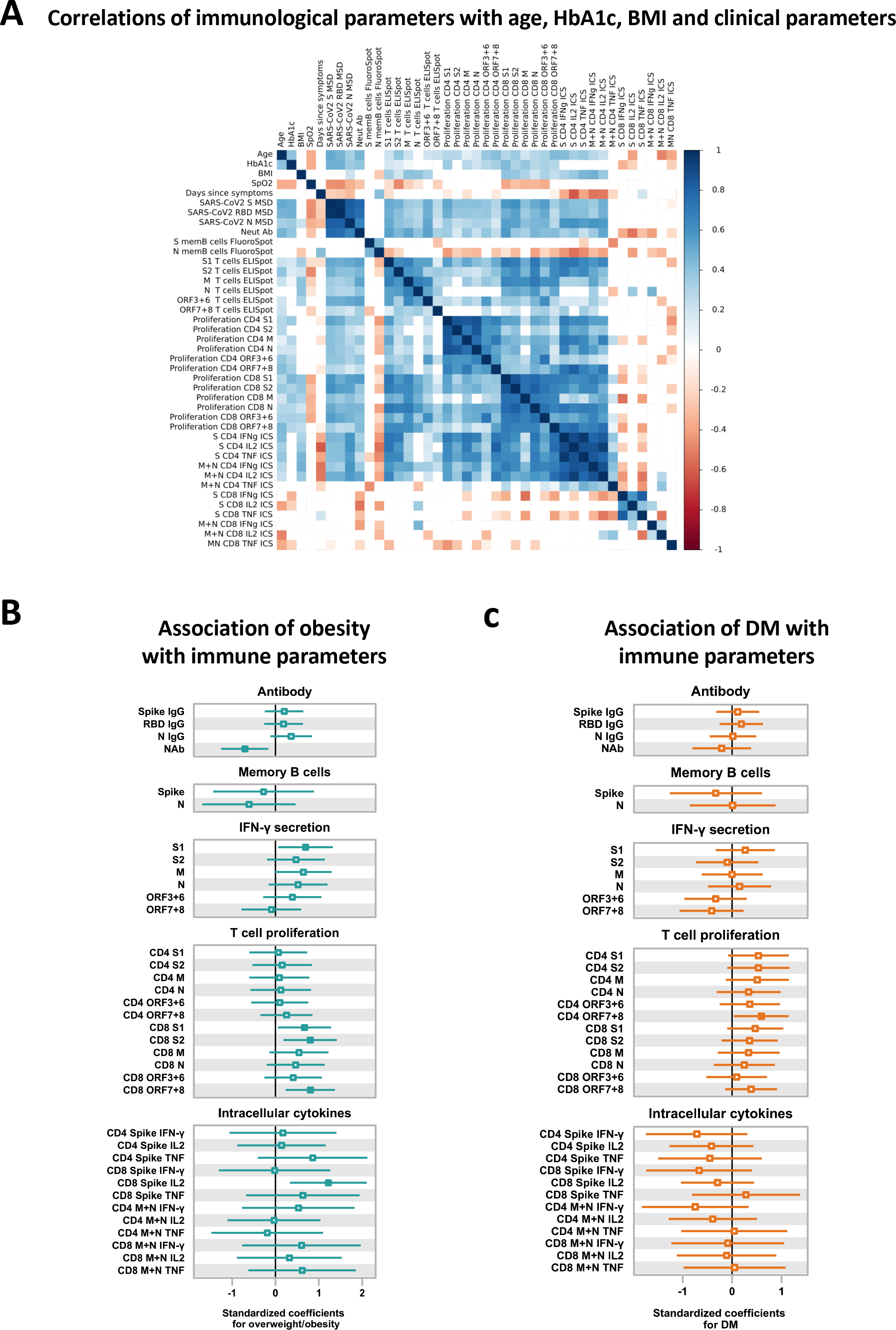
Associations of BMI and HbA1c with antibody, memory B cell, and T cell responses to SARS-CoV-2 (A) Correlations of immunological parameters with age, HbA1c, BMI, SpO2 (lowest recorded oxygen saturation in blood during infection) and days post symptoms onset in SARS-CoV-2 recovered patients. Spearman’s correlation coefficient (colour coded) and only significant values are shown after adjusting for multiple testing using the Benjamini-Hochberg correction at the significance threshold FDR < 0.05. (B) Forest plot illustrating associations of overweight/obesity with antibody, memory B cells, IFN-γ secretion, T cell proliferation, and intracellular cytokine responses to SARS-CoV-2 in recovered patients. The green point estimates represent the standardised unit changes of immunological parameters in overweight/obesity, while adjusted for age, sex, diabetes status, disease severity, and days post symptoms onset. (C) Forest plot illustrating associations of diabetes mellitus with immune responses to SARS-CoV-2 in recovered patients. The red point estimates represent the standardised unit changes of immunological parameters in diabetes, while adjusted for age, sex, obesity status, disease severity and days post symptoms onset. Error bars represent 95% confidence intervals. The non-significant (p ≥ 0.05) results are displayed as hollow points. Detailed results from regression models are shown in table S2. Error bars for (B) and (C) represent 95% confidence intervals, and the non-significant (p ≥ 0.05) results are displayed as hollow points. Detailed results from regression models used for (B) and (C) are shown in table S2.

HbA1c positively correlated with IgG and neutralising antibody titers, CD4+ and CD8+ T cell proliferation, while BMI positively correlated with IFN-γ ELISpot responses, CD8+ T cell proliferation, and CD4 cytokine responses. Notably, we observed inverse relationships between SpO_2_ with IgG and neutralising antibody titers, IFN-γ ELISpot responses, CD8+ T cell proliferative responses, which indicates that more severe disease induces higher antibody and T cell responses following recovery. The time since symptom onset was negatively correlated with the antibody responses and CD4 cytokine responses to SARS-CoV-2.

### Obesity is independently associated with immune parameters in SARS-CoV-2 recovered individuals

Next, we conducted multivariable regression analyses to investigate the individual associations of obesity and diabetes mellitus (DM) on immune responses to SARS-CoV-2 (Figure 5B-C and Tables S2a to S2f). Our analysis, which accounted for age, sex, diabetes status, disease severity and time since symptom onset, revealed that obesity is associated with reduced neutralising antibody responses to SARS-CoV-2 (Figure 5B, Table S2a). In addition, obesity was linked to a higher IFN-γ response to SARS-CoV-2 S1, a higher CD8+ T cells proliferation to S1, S2, ORF 7 and 8, and an increased CD8 IL-2 response to spike (Figure 5B, Tables S2b, S2d and S2f). On the other hand, after adjusting for obesity and other factors, we found no association between DM and antibody or T cell responses to SARS-CoV-2, except a higher proportion of proliferating CD4+ T cells in DM in responses to ORF 7 and 8 (Figure 5C, Table S2a to S2f). We also performed principal component analysis (PCA) to examine the immunological differences in obesity and DM (Figure S5). Our PCA revealed that immunological parameters, including IgG antibody, IFN-y ELISpot, and T cell proliferation, account for 42.7% of the variance among SARS-CoV-2-recovered individuals, and separation of individuals was driven by obesity status (Figure S5A). The key variables in explaining the variability between individuals were anti-N IgG, anti-RBD IgG, anti-S IgG, CD8+ proliferation to N, S1 and S2, and CD4+ proliferation to S1, N and M (Figure S5B-C). Overall, our results indicate diminished neutralising capacity of antibodies and higher T cell responses in obesity, whereas immune responses remain unchanged in DM.

## Discussion

Our study provides insights into the adaptive immunity to SARS-CoV-2 in adults with obesity and diabetes in Bangladesh, where both comorbidities are significant public health concerns. We found that disease severity is associated with higher antibody and T cell responses to SARS-CoV-2 post-recovery, which is consistent with prior studies on severe COVID-19 (33, 34). Also, recovered individuals with symptomatic PCR-confirmed SARS-CoV-2 infection had stronger immune responses compared to asymptomatic seropositive controls, aligning with our previous research in the UK (35), and other studies in Bangladesh (36, 37), and likely attributable to higher antigen exposure in more severe disease.

Our study has revealed a striking disparity in neutralising antibody levels among individuals with overweight/obesity (Ov/Ob) compared to those who are lean. Despite comparable anti-SARS-CoV-2 IgG levels, individuals with Ov/Ob exhibited significantly lower neutralising antibody titers. Notably, this discrepancy persisted even after controlling for age, sex, diabetes status, disease severity, and time since the onset of symptoms. A recent UK cohort study reported rapid waning of neutralising antibodies following COVID-19 vaccination in individuals with severe obesity (38). Another USA cohort study showed lower SARS-CoV-2-specific IgG titers and antibody neutralisation in obesity (39). Our findings highlight the need to investigate the mechanisms underlying the lower neutralising capacity of antibodies in obesity, which could have critical implications for developing effective interventions and treatments for COVID-19 in diverse populations.

T cells play a crucial role in long-term protection against SARS-CoV-2 infection (40), yet there is limited research on the impact of obesity on T cell responses following natural infection. A few studies report that adults with obesity have similar T cell responses to SARS-CoV-2 compared to lean adults after recovery from COVID-19 (41, 42). Nonetheless, these studies did not consider disease severity, a crucial confounder of adaptive immune responses, as shown in our study. We found increased IFN-γ responses to SARS-CoV-2 in Ov/Ob, which were further supported by proliferation assays and ICS showing higher SARS-CoV-2-specific CD8+ T cell proliferation and cytokine responses in Ov/Ob. Moreover, our univariable correlation analyses demonstrate a positive association between higher BMI and increased IFN-γ ELISpot responses, CD8+ T cell proliferation, and CD4 cytokine responses. These findings of higher T cell responses in Ov/Ob persisted in our multivariable regression analyses, which accounts for confounding factors such as age, sex, diabetes status, disease severity, and time components.

In obesity, we hypothesise that the pre-existing immune dysregulation characterised by low-grade chronic inflammation (22, 23), neutrophil activation (43), altered cytokine profile (26), dysregulated T cell homeostasis (44) and impaired cellular immunity (27–29) may lead to delayed viral clearance in SARS-CoV-2 infection. Animal studies have shown increased viral load in obesity (45). Adipose tissue may also serve as a reservoir for the virus (19). Such “depot effect” of prolonged exposure to the virus may contribute to the development of the compensatory higher T cell responses as observed in our individuals with Ov/Ob who survived SARS-CoV-2 infection despite having poor neutralising antibody titers. Further longitudinal studies are needed to understand the underlying mechanisms of T cell immunity in obesity during acute infection and recovery.

The interplay between obesity and diabetes in COVID-19 warrants the need to study the adaptive immunity to SARS-CoV-2 in diabetes. We found that individuals with diabetes had elevated anti-SARS-CoV-2 IgG levels and greater proliferative CD4+ T cell responses. However, after controlling for disease severity, obesity status and other confounding variables, the associations between diabetes and immune responses did not persist. Our analyses suggest that those initially observed associations were potentially driven by age and disease severity. Here we emphasise the critical importance of considering these confounding variables in future studies examining the relationship between DM and COVID- 19 immune responses.

Our study has several limitations. The participants were mostly healthcare workers (HCWs) with a male majority. HCWs self-reported their symptoms, the time of onset, and SpO2 readings during infection, which may introduce recall bias. We could not perform all assays on all participants due to either limited samples or laboratory capacity. We did not evaluate innate immune responses or mucosal immunity, both of which may be impacted by obesity and DM. Intracellular staining revealed low level detectable CD8+ cytokine responses to SARS-CoV-2 in the healthy seronegative controls, suggesting background noise in the assay. In addition, the length of the peptide pools (18-mers) was better optimised for the detection of CD4+ responses than CD8+ responses.

## Conclusion

Our study suggests that obesity is independently associated with lower neutralising antibody levels and higher T cell responses to SARS-CoV-2 following recovery. However, the antiviral adaptive immune responses are preserved in DM. Further analysis using single-cell transcriptomics and flow cytometry will be used to characterise qualitative differences in immune cell subsets.

## Data Availability

All data produced in the present study are available upon reasonable request to the authors

## Acknowledgements

We are grateful to all participants in our study. We sincerely thank Advanced Biomedical Research Laboratory, Department of Biochemistry and Molecular Biology, BSMMU for providing laboratory equipment for sample processing and storage in Bangladesh. We thank the PITCH Consortium, funded by the UK Department of Health and Social Care, for supporting the immunological study in Oxford.

## Author Contributions

MA, PK, FRC, and SJD conceptualised the study. MA, AO, PK, FRC, and SJD developed the methodology. MA, FUHC, AH, NH, and SP were involved in recruitment of the study participants. MA processed the samples. MA carried out the cellular assays. SL, TT, and SLa conducted the antibody binding assays under supervision of MC. PR, AB, SA, HDA, PA, MMR, MMA, SPar, FHM, MMH, SCM, SKB, LT, FRC and AT provided resources. MA curated the data. MA and IN conducted the formal analysis. MA created the data visualisations. MA wrote the original draft. SJD, BK, JH, CPH, PK, IN reviewed and edited the manuscript. BK, AO, PK, FRC and SJD supervised the study. MA, PK, and SJD acquired the funding. AT, LT, TdS, JF, EB, PK and SJD oversaw the data analysis for the study. MA and SJD had unrestricted access to all the data. All authors approved the final draft of the manuscript and take responsibility for its content, including the accuracy of the data.

## References

1. Zhou F, Yu T, Du R, et al. Clinical course and risk factors for mortality of adult inpatients with COVID-19 in Wuhan, China: a retrospective cohort study. The lancet. 2020;395(10229):1054-62.

2. Lighter J, Phillips M, Hochman S, et al. Obesity in patients younger than 60 years is a risk factor for Covid-19 hospital admission. Clinical Infectious Diseases. 2020;71(15):896–7.

3. Palaiodimos L, Kokkinidis DG, Li W, et al. Severe obesity, increasing age and male sex are independently associated with worse in-hospital outcomes, and higher in-hospital mortality, in a cohort of patients with COVID-19 in the Bronx, New York. Metabolism. 2020;108:154262.

4. Hendren NS, De Lemos JA, Ayers C, et al. Association of body mass index and age with morbidity and mortality in patients hospitalized with COVID-19: results from the American Heart Association COVID-19 Cardiovascular Disease Registry. Circulation. 2021;143(2):135–44.

5. Gao M, Piernas C, Astbury NM, et al. Associations between body-mass index and COVID-19 severity in 6·9 million people in England: a prospective, community-based, cohort study. The Lancet Diabetes & Endocrinology. 2021;9(6):350–9.

6. Huttunen R, Syrjänen J. Obesity and the risk and outcome of infection. International journal of obesity. 2013;37(3):333–40.

7. Benfield T, Jensen JS, Nordestgaard BG. Influence of diabetes and hyperglycaemia on infectious disease hospitalisation and outcome. Diabetologia. 2007;50(3):549–54.

8. Muller LM, Gorter KJ, Hak E, et al. Increased risk of common infections in patients with type 1 and type 2 diabetes mellitus. Clin Infect Dis. 2005;41(3):281–8.

9. Dunachie S, Chamnan P. The double burden of diabetes and global infection in low and middle-income countries. Trans R Soc Trop Med Hyg. 2019;113(2):56–64.

10. Reilly SM, Saltiel AR. Adapting to obesity with adipose tissue inflammation. Nature Reviews Endocrinology. 2017;13(11):633–43.

11. Roberts AC, Porter KE. Cellular and molecular mechanisms of endothelial dysfunction in diabetes. Diabetes and Vascular Disease Research2013.

12. Klöting N, Blüher M. Adipocyte dysfunction, inflammation and metabolic syndrome. Reviews in Endocrine and Metabolic Disorders. 2014;15(4):277–87.

13. Louwen F, Ritter A, Kreis N, Yuan J. Insight into the development of obesity: functional alterations of adipose-derived mesenchymal stem cells. Obesity Reviews. 2018;19(7):888–904.

14. Febbraio MA. Role of interleukins in obesity: implications for metabolic disease. Trends in Endocrinology & Metabolism. 2014;25(6):312–9.

15. McLaughlin T, Ackerman SE, Shen L, Engleman E. Role of innate and adaptive immunity in obesity-associated metabolic disease. The Journal of clinical investigation. 2017;127(1):5–13.

16. Hodgson K, Morris J, Bridson T, Govan B, Rush C, Ketheesan N. Immunological mechanisms contributing to the double burden of diabetes and intracellular bacterial infections. Immunology. 2015;144(2):171–85.

17. Lim S, Bae JH, Kwon H-S, Nauck MA. COVID-19 and diabetes mellitus: from pathophysiology to clinical management. Nature Reviews Endocrinology. 2021;17(1):11–30.

18. Kruglikov IL, Scherer PE. The role of adipocytes and adipocyte-like cells in the severity of COVID-19 infections. Obesity. 2020;28(7):1187–90.

19. Martinez-Colon GJ, Ratnasiri K, Chen H, et al. SARS-CoV-2 infection drives an inflammatory response in human adipose tissue through infection of adipocytes and macrophages. Sci Transl Med. 2022;14(674):eabm9151.

20. Office for National Statistics, UK. Updating ethnic contrasts in deaths involving the coronavirus (COVID-19), England: 10 January 2022 to 16 February 2022. Updated April 7, 2022. Accessed April 3, 2023. https://www.ons.gov.uk/peoplepopulationandcommunity/birthsdeathsandmarriages/deaths/articles/updatingethniccontrastsindeathsinvolvingthecoronaviruscovid19englandandwales/10january2022to16february2022.

21. Office for National Statistics, UK. Updating ethnic contrasts in deaths involving the coronavirus (COVID-19), England: 24 January 2020 to 31 March 2021. Updated May 26, 2021, Accessed October 21, 2022. https://www.ons.gov.uk/peoplepopulationandcommunity/birthsdeathsandmarriages/deaths/articles/updatingethniccontrastsindeathsinvolvingthecoronaviruscovid19englandandwales/24january2020to31march2021.

22. Siddiq S, Ahmed S, Akram I. Clinical outcomes following COVID-19 infection in ethnic minority groups in the UK: a systematic review and meta-analysis. Public Health. 2022.

23. Gao M, Piernas C, Astbury NM, et al. Associations between body-mass index and COVID-19 severity in 6·9 million people in England: a prospective, community-based, cohort study. The Lancet Diabetes & Endocrinology. 2021;9(6):350–9.

24. Yates T, Zaccardi F, Islam N, et al. Obesity, ethnicity, and risk of critical care, mechanical ventilation, and mortality in patients admitted to hospital with COVID-19: analysis of the ISARIC CCP-UK Cohort. Obesity. 2021;29(7):1223–30.

25. Yates T, Summerfield A, Razieh C, et al. A population-based cohort study of obesity, ethnicity and COVID-19 mortality in 12.6 million adults in England. Nature Communications 2022 13:1. 2022;13(1):1-9.

26. Akhtar S, Nasir JA, Sarwar A, et al. Prevalence of diabetes and pre-diabetes in Bangladesh: a systematic review and meta-analysis. BMJ open. 2020;10(9):e036086.

27. Ogbe A, Kronsteiner B, Skelly DT, et al. T cell assays differentiate clinical and subclinical SARS-CoV-2 infections from cross-reactive antiviral responses. Nat Commun. 2021;12(1):2055.

28. Angyal A, Longet S, Moore SC, et al. T-cell and antibody responses to first BNT162b2 vaccine dose in previously infected and SARS-CoV-2-naive UK health-care workers: a multicentre prospective cohort study. Lancet Microbe. 2022;3(1):e21–e31.

29. Huang KA, Tan TK, Chen TH, et al. Breadth and function of antibody response to acute SARS-CoV-2 infection in humans. PLoS Pathog. 2021;17(2):e1009352.

30. Moore SC, Kronsteiner B, Longet S, et al. Evolution of long-term vaccine-induced and hybrid immunity in healthcare workers after different COVID-19 vaccine regimens. Med. 2023;4(3):191–215 e9.

31. Tomic A, Tomic I, Waldron L, et al. SIMON: Open-Source Knowledge Discovery Platform. Patterns. 2021;2(1):100178.

32. Consultation WE. Appropriate body-mass index for Asian populations and its implications for policy and intervention strategies. Lancet (London, England). 2004;363(9403):157-63.

33. Lynch KL, Whitman JD, Lacanienta NP, et al. Magnitude and Kinetics of Anti-Severe Acute Respiratory Syndrome Coronavirus 2 Antibody Responses and Their Relationship to Disease Severity. Clin Infect Dis. 2021;72(2):301–8.

34. Peng Y, Mentzer AJ, Liu G, et al. Broad and strong memory CD4(+) and CD8(+) T cells induced by SARS-CoV-2 in UK convalescent individuals following COVID-19. Nat Immunol. 2020;21(11):1336–45.

35. Tomic A, Skelly DT, Ogbe A, et al. Divergent trajectories of antiviral memory after SARS-CoV- 2 infection. Nature Communications. 2022;13(1):1251.

36. Shirin T, Bhuiyan TR, Charles RC, et al. Antibody responses after COVID-19 infection in patients who are mildly symptomatic or asymptomatic in Bangladesh. Int J Infect Dis. 2020;101:220–5.

37. Bhuiyan TR, Al Banna H, Kaisar MH, et al. Correlation of antigen-specific immune response with disease severity among COVID-19 patients in Bangladesh. Frontiers in Immunology. 2022;13.

38. van der Klaauw AA, Horner EC, Pereyra-Gerber P, et al. Accelerated waning of the humoral response to COVID-19 vaccines in obesity. Nature Medicine. 2023.

39. Frasca D, Reidy L, Romero M, et al. The majority of SARS-CoV-2-specific antibodies in COVID- 19 patients with obesity are autoimmune and not neutralizing. International Journal of Obesity. 2022;46(2):427–32.

40. Feng C, Shi J, Fan Q, et al. Protective humoral and cellular immune responses to SARS-CoV-2 persist up to 1 year after recovery. Nature communications. 2021;12(1):1–7.

41. Nilles EJ, Siddiqui SM, Fischinger S, et al. Epidemiological and immunological features of obesity and SARS-CoV-2. Viruses. 2021;13(11):2235.

42. Wrigley Kelly NE, Kenny G, Cassidy FC, et al. Individuals with obesity who survive SARS-CoV-2 infection have preserved antigen specific T cell frequencies. Obesity. 2022.

43. Ali M, Jasmin S, Fariduddin M, Alam SMK, Arslan MI, Biswas SK. Neutrophil elastase and myeloperoxidase mRNA expression in overweight and obese subjects. Molecular Biology Reports. 2018;45(5):1245–52.

44. Paich HA, Sheridan PA, Handy J, et al. Overweight and obese adult humans have a defective cellular immune response to pandemic H1N1 influenza A virus. Obesity. 2013;21(11):2377–86.

45. Lee KS, Russ BP, Wong TY, et al. Obesity and metabolic dysfunction drive sex-associated differential disease profiles in hACE2-mice challenged with SARS-CoV-2. iScience. 2022;25(10):105038.

