## Supporting information for "Obesity Differs from Diabetes Mellitus in Antibody and T Cell Responses Post COVID-19 Recovery"

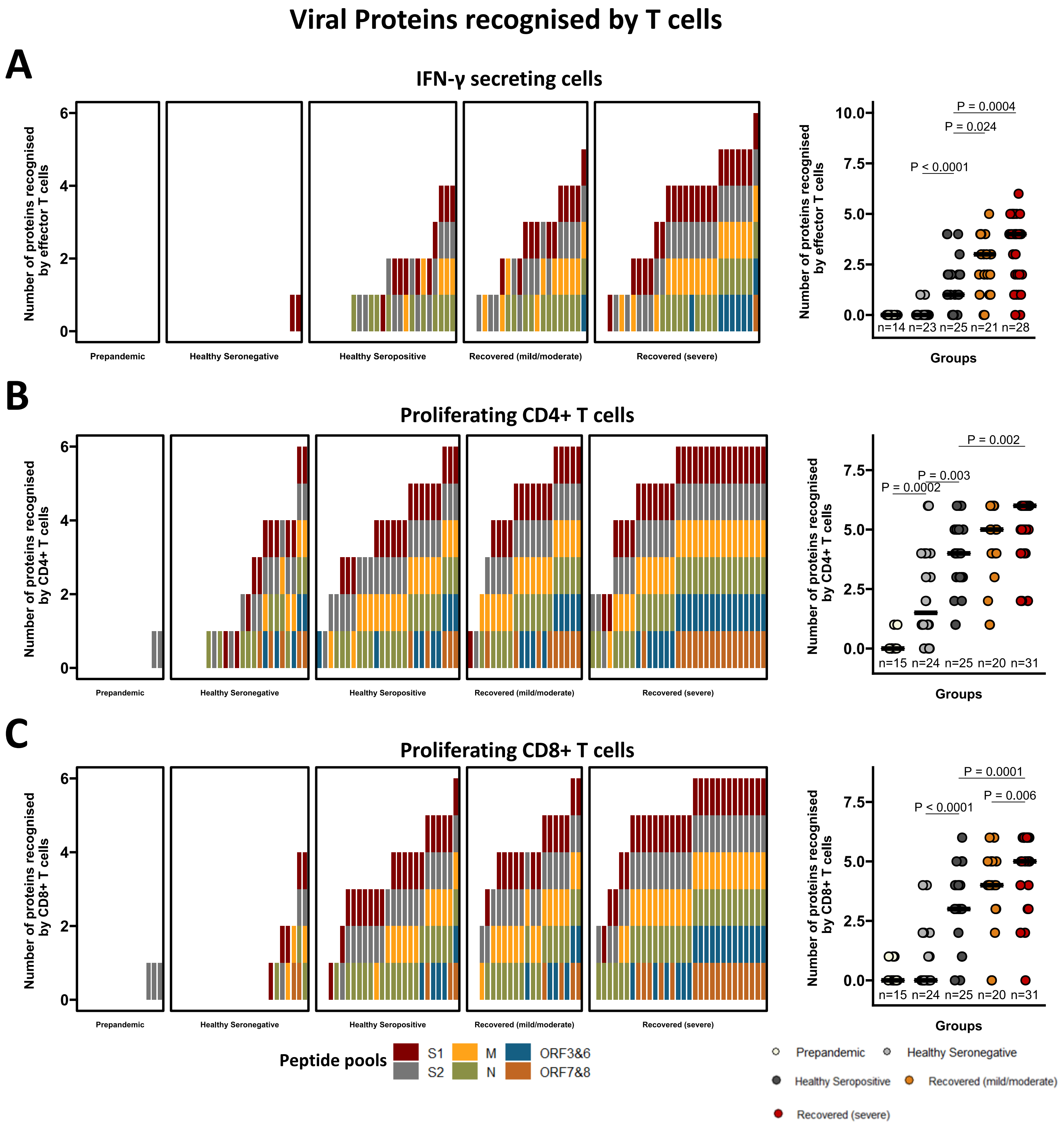


**Figure S1: The breadth of T cell responses to SARS-CoV-2 was higher in patients who recovered from severe COVID-19.**

Barplots on the left display the number of viral proteins recognised in individuals across groups (coloured by specificity) by (A) ex vivo IFN-γ ELISpot assay, (B) CD4+ T cell proliferation, (C) CD8+ T cell proliferation. Dot plots on the right show the comparison of the number of viral proteins recognised among groups by corresponding assays.

A two-tailed Wilcoxon rank-sum test was used to compare between the groups (without correction for multiple testing), p values are shown on the top of the dot plots The number of individuals (n) evaluated per assay is displayed at the bottom of the corresponding dot plots, while horizontal bars represent the medians.

**A** **Gating strategy for T cell proliferation**

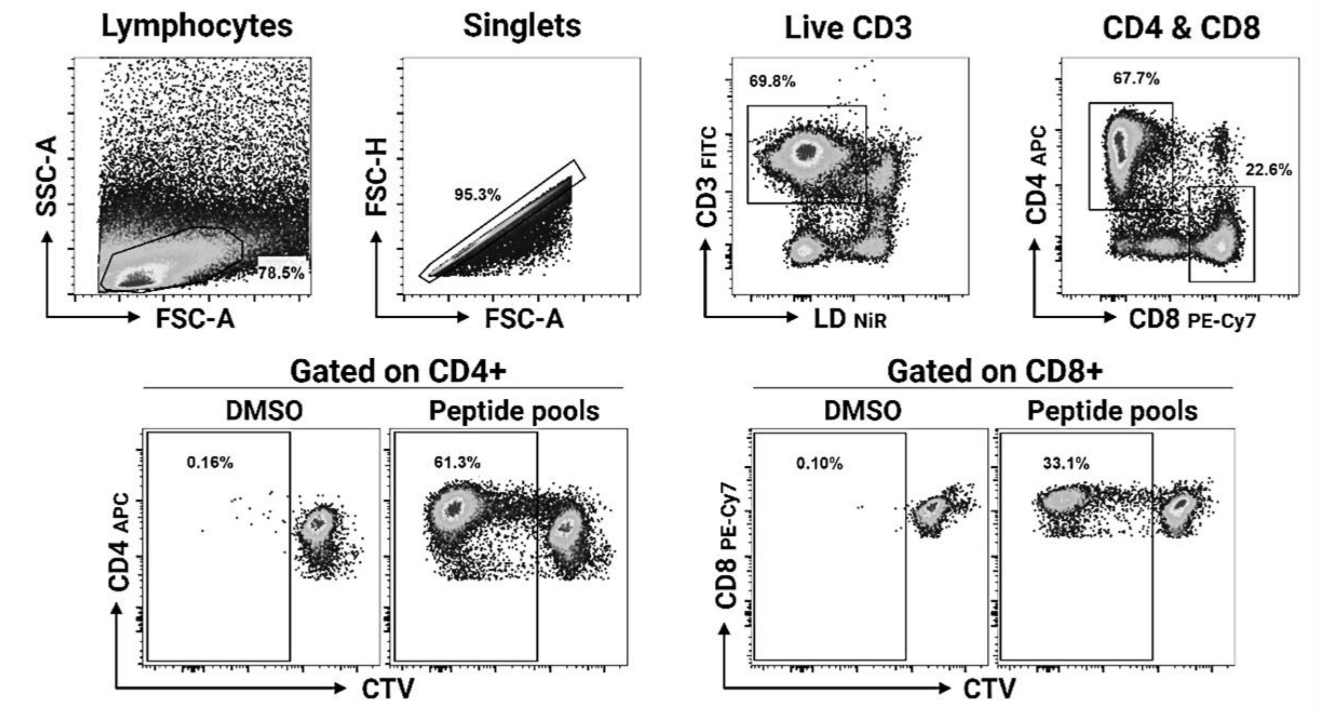


**B** **Gating strategy for antigen-specific T cell cytokine secretion (ICS)**

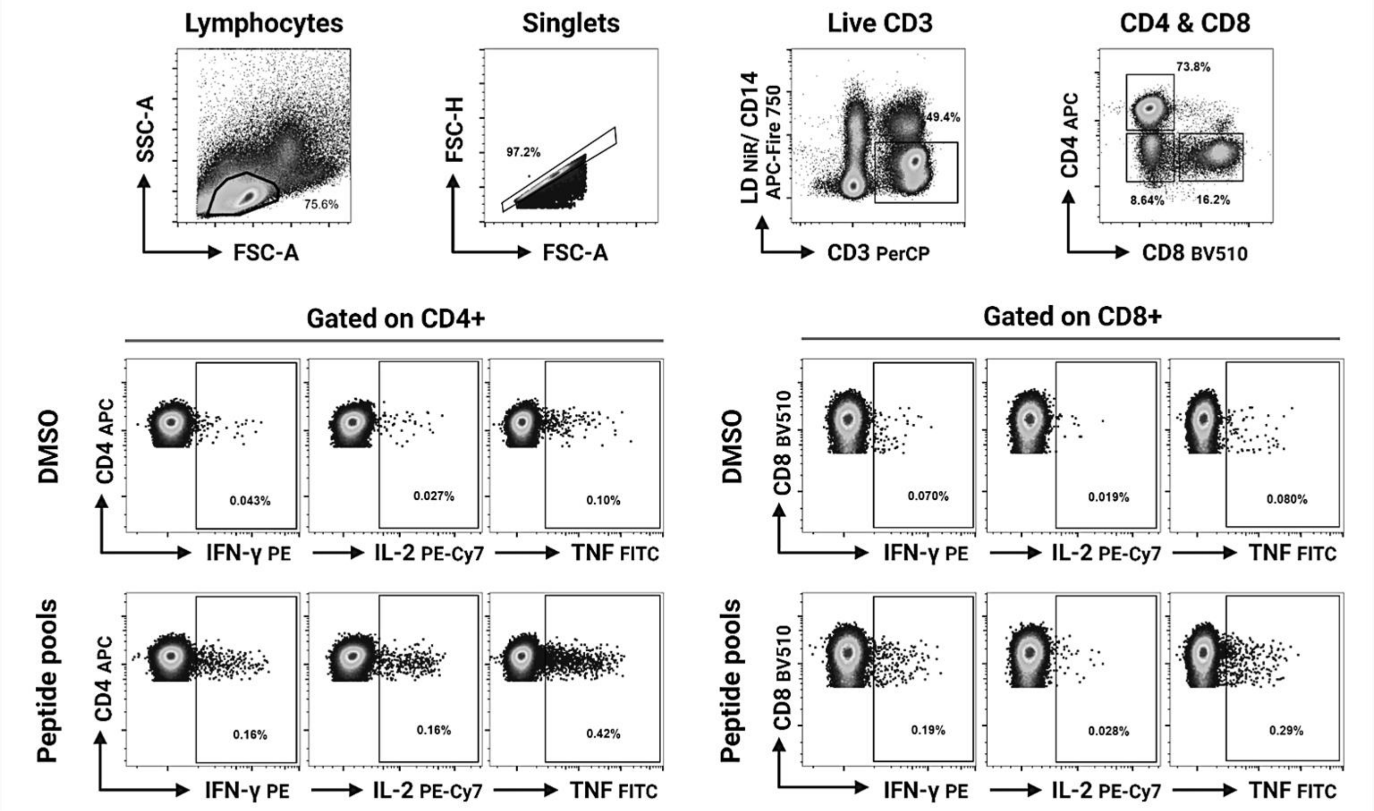

#### Figure S2: Gating strategy for T cell proliferation and intracellular cytokine staining (ICS).

(A) For T cell proliferation assays, lymphocytes were gated using forward and side scatter area (FSC-A and SSC-A) parameters, followed by single cell gating on FSC-height (H) and area (A). Live T cells were then gated (LD-NiR low CD3+) and T cell subsets were identified (CD4+CD8- and CD8+CD4-) using CD4+ APC and CD8+ PE-Cy7. Within the CD4+ and the CD8+ T cell gate proliferating cells were identified by gating on cells with reduced CTV (CellTraceTM Violet) fluorescence intensity.

(B) For ICS assays lymphocytes were gated using forward and side scatter area (FSC-A and SSC-A) parameters, followed by single cell gating on FSC-height (H) and area (A). Live CD3+ T cells were gated based on exclusion of dead cells (LD-NiR) and monocytes (CD14 APC Fire-750) as well as positivity for CD3 PerCP. T cell subsets were identified based on staining for CD4 APC and CD8 BV510 respectively and expression of cytokines (IFN-γ, TNF, IL-2) was then identified in the CD4+CD8- gate as well as the CD8+CD4- gate. Representative gating is shown for the DMSO negative control and the reactive peptide pools.

A

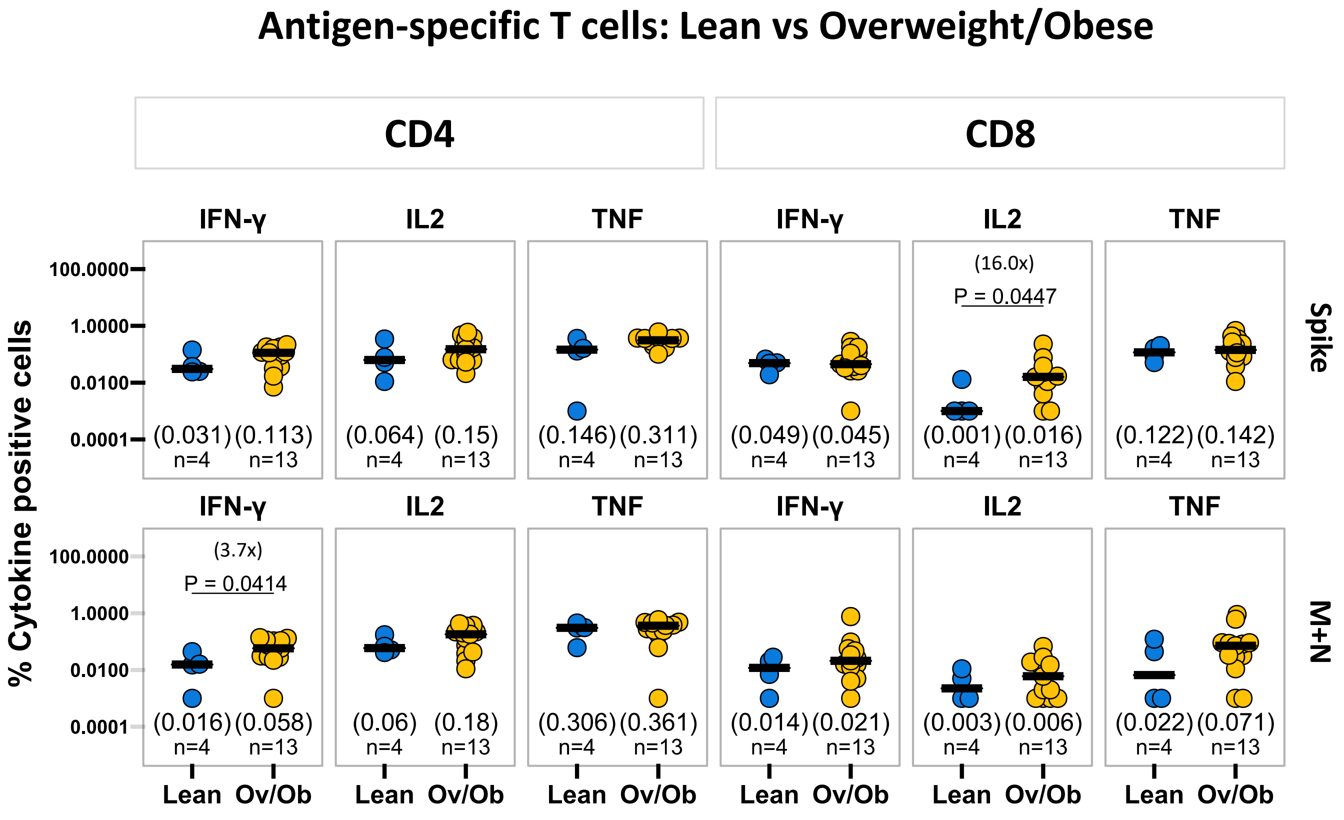

Antigen-specific T cells: Lean vs Overweight/Obese
CD4
CD8
IFN-γ
IL2
TNF
% Cytokine positive cells
100.0000
1.0000
0.0100
0.0001
Spike
M+N
(16.0x)
P = 0.0447
(3.7x)
P = 0.0414
(0.031) (0.113)
(0.064) (0.15)
(0.146) (0.311)
(0.049) (0.045)
(0.001) (0.016)
(0.122) (0.142)
(0.016) (0.058)
(0.06) (0.18)
(0.306) (0.361)
(0.014) (0.021)
(0.003) (0.006)
(0.022) (0.071)
n=4 n=13
Lean Ov/Ob


B

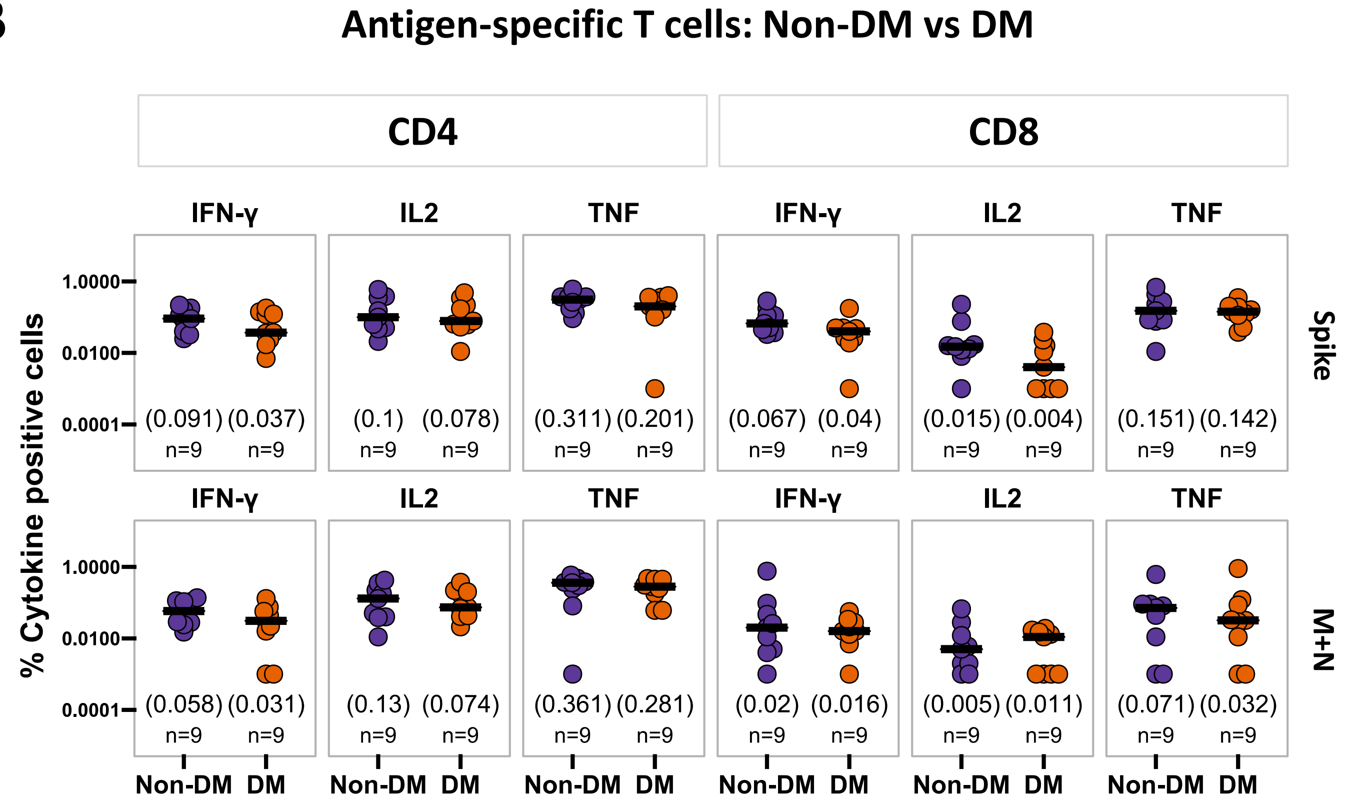

Antigen-specific T cells: Non-DM vs DM
CD4
CD8
IFN-γ
IL2
TNF
% Cytokine positive cells
1.0000
0.0100
0.0001
Spike
M+N
(0.091) (0.037)
(0.1) (0.078)
(0.311) (0.201)
(0.067) (0.04)
(0.015) (0.004)
(0.151) (0.142)
(0.058) (0.031)
(0.13) (0.074)
(0.361) (0.281)
(0.02) (0.016)
(0.005) (0.011)
(0.071) (0.032)
n=9 n=9
Non-DM DM

**Figure S3: Overweight/obese status is associated with increased T cell cytokine responses to SARS-CoV-2 after recovery, but the cytokine responses are similar between diabetic and non-diabetic recovered patients.**

Comparison of cytokine responses (IFN-γ, IL-2 and TNF) by $CD4^+$ and $CD8^+$ T cells in response to SARS-CoV2 spike (top panel) and M+N pools (bottom panel) between (A) lean and overweight/obese (Ov/Ob), (B) non-diabetic (Non-DM) and diabetic (DM) recovered COVID-19 patients.

A two-tailed Wilcoxon rank-sum test was used to compare between the groups (without correction for multiple testing), with fold changes in brackets. P values are displayed in case of significant differences. The number of individuals (n) evaluated per assay is displayed at the bottom of the corresponding dot plots. Horizontal bars represent the medians, and the median values are shown in brackets immediately above the number of individuals (n) in each column.

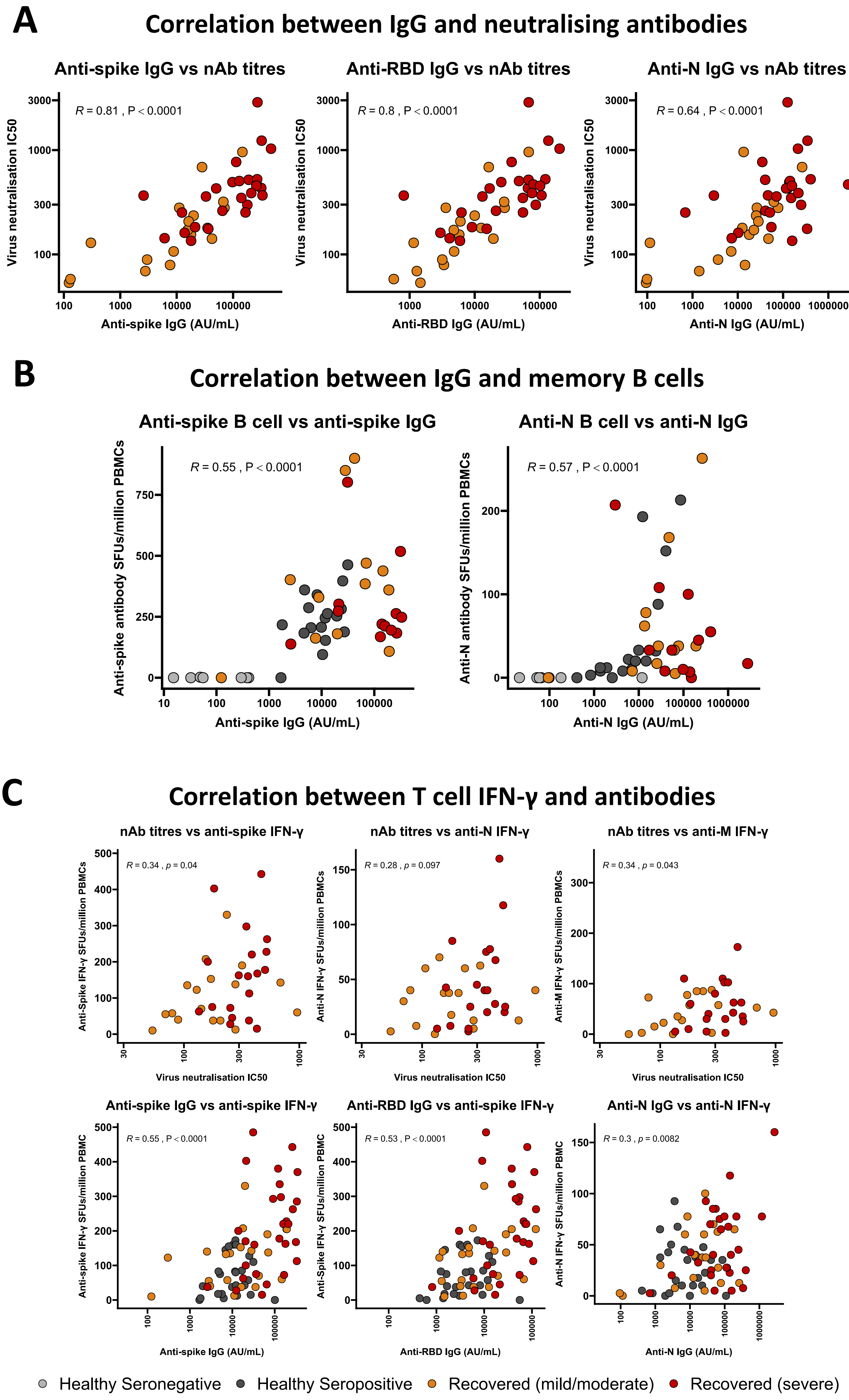
A
Correlation between IgG and neutralising antibodies
Anti-spike IgG vs nAb titres
R = 0.81 , P < 0.0001
Virus neutralisation IC50
Anti-spike IgG (AU/mL)
Anti-RBD IgG vs nAb titres
R = 0.8 , P < 0.0001
Anti-RBD IgG (AU/mL)
Anti-N IgG vs nAb titres
R = 0.64 , P < 0.0001
Anti-N IgG (AU/mL)
B
Correlation between IgG and memory B cells
Anti-spike B cell vs anti-spike IgG
R = 0.55 , P < 0.0001
Anti-spike antibody SFUs/million PBMCs
Anti-spike IgG (AU/mL)
Anti-N B cell vs anti-N IgG
R = 0.57 , P < 0.0001
Anti-N antibody SFUs/million PBMCs
Anti-N IgG (AU/mL)
C
Correlation between T cell IFN-γ and antibodies
nAb titres vs anti-spike IFN-γ
R = 0.34 , p = 0.04
Anti-Spike IFN-γ SFUs/million PBMCs
Virus neutralisation IC50
nAb titres vs anti-N IFN-γ
R = 0.28 , p = 0.097
Anti-N IFN-γ SFUs/million PBMCs
nAb titres vs anti-M IFN-γ
R = 0.34 , p = 0.043
Anti-M IFN-γ SFUs/million PBMCs
Anti-spike IgG vs anti-spike IFN-γ
R = 0.55 , P < 0.0001
Anti-spike IFN-γ SFUs/million PBMC
Anti-spike IgG (AU/mL)
Anti-RBD IgG vs anti-spike IFN-γ
R = 0.53 , P < 0.0001
Anti-Spike IFN-γ SFUs/million PBMC
Anti-RBD IgG (AU/mL)
Anti-N IgG vs anti-N IFN-γ
R = 0.3 , p = 0.0082
Anti-N IFN-γ SFUs/million PBMC
Anti-N IgG (AU/mL)
Healthy Seronegative
Healthy Seropositive
Recovered (mild/moderate)
Recovered (severe)

**Figure S4: Correlation between antibody, B cell and T cell responses to SARS-CoV-2.**

(A) Correlation of neutralising antibody (nAb) titers with IgG responses to SARS-CoV-2 spike, receptor binding domain (RBD), and nucleoprotein (N) in recovered COVID-19 patients.

(B) Correlation between IgG and memory B cell responses to SARS-CoV-2 spike (left) and N (right) in healthy seronegative and seropositive controls and recovered patients.

(C) Correlation of IFN-γ ELISpot responses with IgG and nAb titers in healthy seropositive controls and recovered patients.

Spearman's correlation coefficient (R) and p-values are shown at the top of each plot.

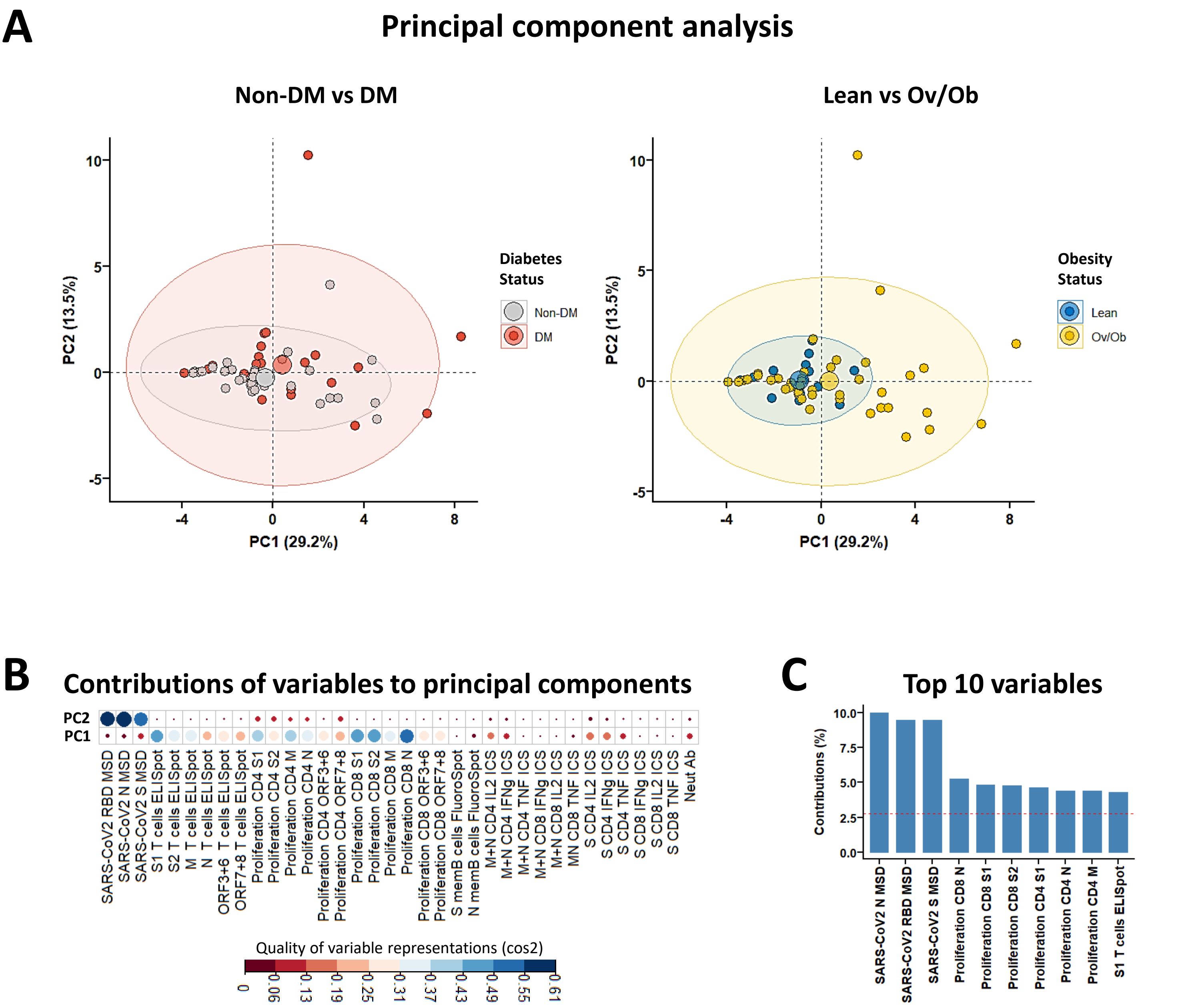


**Figure S5: Principal component analysis with integrated immunological data in recovered COVID-19 patients.**

(A) PCA plot representing integrated immunological data, grouped based on the diabetes (left) and obesity (right) status. Percentage indicates the variance explained by the principal component (PC).

(B) Quality of variable representations (colour-coded, cos2) and contributions of variables to principal components 1 and 2 (size of the circle).

(C) Top 10 variables and their contribution to PC 1 and 2.

**Table S1a: Demographic characteristics of recovered participants with or without obesity.**

| **Variable** | **Normal weight** (N = 20) | **Overweight/obesity** (N = 52) | **p-value** |
|---|---|---|---|
| **Age in years**, median (IQR[#]) | 55 (43, 61) | 43 (34, 53) | 0.020 [a] |
| **Sex** | | | >0.9 [b] |
| Female, n (%) | 3 (15%) | 8 (15%) | |
| Male, n (%) | 17 (85%) | 44 (85%) | |
| **Diabetes status** | | | 0.082 [b] |
| Non-diabetic, n (%) | 9 (45%) | 35 (67%) | |
| Diabetic, n (%) | 11 (55%) | 17 (33%) | |
| **Disease severity** | | | >0.9 [b] |
| Mild/moderate, n (%) | 6 (30%) | 22 (42%) | |
| Severe, n (%) | 14 (70%) | 30 (58%) | |
| **Days post symptom onset**, median (IQR[#]) | 54 (36, 110) | 59 (38, 100) | >0.9 [a] |

[#]IQR (Interquartile range)
[a] Wilcoxon rank sum test; [b] Pearson's Chi-squared test

**Table S1b: Demographic characteristics of recovered participants with or without diabetes.**

| **Variable** | **Non-diabetic** (N = 46) | **Diabetic** (N = 29) | **p-value** |
|---|---|---|---|
| **Age in years**, median (IQR#) | 40 (31, 51) | 53 (45, 63) | <0.001 [a] |
| **Sex** | | | >0.9 [b] |
| Female, n (%) | 7 (15%) | 4 (14%) | |
| Male, n (%) | 39 (85%) | 25 (86%) | |
| **Obesity category** | | | 0.21 [b] |
| Normal weight, n (%) | 9 (20%) | 11 (38%) | |
| Overweight/obesity, n (%) | 35 (76%) | 17 (59%) | |
| Data missing, n (%) | 2 (4%) | 1 (3%) | |
| **Disease severity** | | | 0.027 [b] |
| Mild/moderate, n (%) | 24 (52%) | 7 (24%) | |
| Severe, n (%) | 22 (48%) | 22 (76%) | |
| **Days post symptom onset**, median (IQR#) | 69 (33, 120) | 52 (40, 70) | 0.3 [a] |

#IQR (Interquartile range)
[a] Wilcoxon rank sum test; [b] Pearson's Chi-squared test

**Table S2a. Generalised linear regression models of antibody (IgG, neutralising Ab) and memory B cell responses in SARS-CoV-2 recovered patients**

| | Anti-spike IgG | | | Anti-RBD IgG | | | Anti-N IgG | | | Neutralising antibody | | | Anti-spike memory B cells | | | Anti-N memory B cells | | |
|---|---|---|---|---|---|---|---|---|---|---|---|---|---|---|---|---|---|---|
| **Coefficient** | **Est** | **SE** | **p value** | **Est** | **SE** | **p value** | **Est** | **SE** | **p value** | **Est** | **SE** | **p value** | **Est** | **SE** | **p value** | **Est** | **SE** | **p value** |
| **Age** | 0.2615 | 0.1316 | 0.0512 | 0.2219 | 0.1319 | 0.0974 | 0.1134 | 0.1421 | 0.4278 | 0.2088 | 0.1794 | 0.2518 | 0.2054 | 0.2667 | 0.4523 | 0.1126 | 0.2476 | 0.6553 |
| **Sex (male)** | -0.2075 | 0.2742 | 0.4519 | -0.1770 | 0.2748 | 0.5219 | -0.1592 | 0.2960 | 0.5925 | -0.1880 | 0.3405 | 0.5843 | 0.2552 | 0.5673 | 0.6589 | -0.0994 | 0.5266 | 0.8526 |
| **DM (yes)** | 0.1133 | 0.2226 | 0.6126 | 0.1848 | 0.2231 | 0.4105 | 0.0159 | 0.2403 | 0.9474 | -0.2130 | 0.3030 | 0.4864 | -0.3309 | 0.4765 | 0.4973 | 0.0078 | 0.4423 | 0.9862 |
| **Obesity (yes)** | 0.1981 | 0.2264 | 0.3846 | 0.1861 | 0.2269 | 0.4152 | 0.3610 | 0.2444 | 0.1445 | -0.7077 | 0.2788 | **0.0155** | -0.2757 | 0.5925 | 0.6481 | -0.6133 | 0.5501 | 0.2813 |
| **Disease Severity (severe)** | 0.7628 | 0.2464 | **0.0029** | 0.7212 | 0.2470 | **0.0048** | 0.8410 | 0.2660 | **0.0024** | 0.6099 | 0.3077 | 0.0549 | 0.1470 | 0.5161 | 0.7794 | -0.0677 | 0.4791 | 0.8895 |
| **Days pso** | -0.1537 | 0.1050 | 0.1482 | -0.2229 | 0.1053 | **0.0381** | -0.1740 | 0.1134 | 0.1296 | -0.1988 | 0.1424 | 0.1711 | 0.2341 | 0.2735 | 0.4047 | 0.3316 | 0.2539 | 0.2100 |

Est (estimates), SE (standard error), pso (post symptom onset)

#### Table S2b. Generalised linear regression models of IFN-γ T cell ELISpot responses in SARS-CoV-2 recovered patients

| | S1 IFN-γ ELISpot | | | S2 IFN-γ ELISpot | | | M IFN-γ ELISpot | | | N IFN-γ ELISpot | | | ORF3+6 IFN-γ ELISpot | | | ORF7+8 IFN-γ ELISpot | | |
|---|---|---|---|---|---|---|---|---|---|---|---|---|---|---|---|---|---|---|
| **Coefficient** | **Est** | **SE** | **p value** | **Est** | **SE** | **p value** | **Est** | **SE** | **p value** | **Est** | **SE** | **p value** | **Est** | **SE** | **p value** | **Est** | **SE** | **p value** |
| **Age** | 0.2439 | 0.1633 | 0.1430 | 0.3061 | 0.1728 | 0.0839 | 0.1504 | 0.1691 | 0.3788 | 0.1408 | 0.1752 | 0.4263 | 0.1169 | 0.1730 | 0.5031 | 0.1425 | 0.1779 | 0.4275 |
| **Sex (male)** | -0.2320 | 0.3547 | 0.5168 | 0.0767 | 0.3752 | 0.8391 | -0.2778 | 0.3672 | 0.4536 | -0.2894 | 0.3806 | 0.4513 | -0.3771 | 0.3758 | 0.3215 | -0.0962 | 0.3863 | 0.8046 |
| **DM (yes)** | 0.2647 | 0.3047 | 0.3900 | -0.1007 | 0.3223 | 0.7563 | -0.0012 | 0.3154 | 0.9971 | 0.1480 | 0.3269 | 0.6532 | -0.3354 | 0.3228 | 0.3049 | -0.4161 | 0.3319 | 0.2170 |
| **Obesity (yes)** | 0.6928 | 0.3229 | **0.0379** | 0.4721 | 0.3416 | 0.1745 | 0.6411 | 0.3343 | 0.0621 | 0.5200 | 0.3465 | 0.1410 | 0.3884 | 0.3421 | 0.2628 | -0.0978 | 0.3517 | 0.7823 |
| **Disease Severity (severe)** | 0.3741 | 0.3112 | 0.2362 | 0.2880 | 0.3292 | 0.3867 | 0.4061 | 0.3221 | 0.2145 | 0.2215 | 0.3339 | 0.5107 | 0.4818 | 0.3297 | 0.1515 | 0.4011 | 0.3389 | 0.2435 |
| **Days pso** | -0.1033 | 0.1446 | 0.4791 | 0.0355 | 0.1530 | 0.8179 | 0.1950 | 0.1497 | 0.2001 | 0.2164 | 0.1552 | 0.1706 | -0.1146 | 0.1532 | 0.4589 | 0.0359 | 0.1575 | 0.8207 |

Est (estimates), SE (standard error), pso (post symptom onset)

#### Table S2c. Generalised linear regression models of CD4 T cell proliferative responses in SARS-CoV-2 recovered patients

| | S1 CD4 proliferation | | | S2 CD4 proliferation | | | M CD4 proliferation | | | N CD4 proliferation | | | ORF3+6 CD4 proliferation | | | ORF7+8 CD4 proliferation | | |
|---|---|---|---|---|---|---|---|---|---|---|---|---|---|---|---|---|---|---|
| **Coefficient** | **Est** | **SE** | **p value** | **Est** | **SE** | **p value** | **Est** | **SE** | **p value** | **Est** | **SE** | **p value** | **Est** | **SE** | **p value** | **Est** | **SE** | **p value** |
| **Age** | 0.2340 | 0.1598 | 0.1504 | 0.2142 | 0.1637 | 0.1978 | 0.0762 | 0.1660 | 0.6486 | 0.1505 | 0.1671 | 0.3727 | 0.3135 | 0.1580 | 0.0536 | 0.4431 | 0.1437 | **0.0036** |
| **Sex (male)** | -0.2905 | 0.3594 | 0.4235 | -0.1127 | 0.3682 | 0.7610 | -0.2992 | 0.3734 | 0.4274 | -0.2926 | 0.3758 | 0.4405 | 0.1262 | 0.3553 | 0.7242 | 0.5546 | 0.3231 | 0.0933 |
| **DM (yes)** | 0.5313 | 0.3147 | 0.0986 | 0.5313 | 0.3224 | 0.1067 | 0.5095 | 0.3270 | 0.1266 | 0.3326 | 0.3291 | 0.3178 | 0.3569 | 0.3112 | 0.2577 | 0.5901 | 0.2830 | **0.0430** |
| **Obesity (yes)** | 0.0663 | 0.3406 | 0.8465 | 0.1532 | 0.3489 | 0.6627 | 0.0884 | 0.3539 | 0.8039 | 0.1225 | 0.3561 | 0.7325 | 0.0989 | 0.3367 | 0.7703 | 0.2534 | 0.3062 | 0.4124 |
| **Disease Severity (severe)** | -0.0093 | 0.3172 | 0.9767 | -0.0542 | 0.3249 | 0.8684 | -0.0908 | 0.3296 | 0.7842 | 0.0633 | 0.3317 | 0.8496 | 0.2303 | 0.3136 | 0.4668 | -0.0186 | 0.2852 | 0.9483 |
| **Days pso** | 0.0012 | 0.1534 | 0.9936 | 0.0623 | 0.1571 | 0.6935 | -0.0291 | 0.1593 | 0.8558 | 0.0033 | 0.1603 | 0.9839 | 0.0507 | 0.1516 | 0.7398 | 0.0141 | 0.1379 | 0.9193 |

Est (estimates), SE (standard error), pso (post symptom onset)

#### Table S2d. Generalised linear regression models of CD8 T cell proliferative responses in SARS-CoV-2 recovered patients

| | S1 CD8 proliferation | | | S2 CD8 proliferation | | | M CD8 proliferation | | | N CD8 proliferation | | | ORF3+6 CD8 proliferation | | | ORF7+8 CD8 proliferation | | |
|---|---|---|---|---|---|---|---|---|---|---|---|---|---|---|---|---|---|---|
| **Coefficient** | **Est** | **SE** | **p value** | **Est** | **SE** | **p value** | **Est** | **SE** | **p value** | **Est** | **SE** | **p value** | **Est** | **SE** | **p value** | **Est** | **SE** | **p value** |
| **Age** | 0.2227 | 0.1469 | 0.1368 | 0.2834 | 0.1473 | 0.0611 | 0.1215 | 0.1627 | 0.4593 | 0.1715 | 0.1595 | 0.2881 | 0.2439 | 0.1588 | 0.1320 | 0.4568 | 0.1356 | **0.0016** |
| **Sex (male)** | -0.3662 | 0.3304 | 0.2738 | -0.4932 | 0.3314 | 0.1440 | -0.3724 | 0.3659 | 0.3145 | -0.1622 | 0.3587 | 0.6534 | -0.2693 | 0.3573 | 0.4551 | 0.1134 | 0.3049 | 0.7118 |
| **DM (yes)** | 0.4680 | 0.2893 | 0.1131 | 0.3532 | 0.2902 | 0.2301 | 0.3330 | 0.3204 | 0.3044 | 0.2438 | 0.3141 | 0.4419 | 0.0904 | 0.3128 | 0.7740 | 0.3822 | 0.2670 | 0.1595 |
| **Obesity (yes)** | 0.6695 | 0.3131 | **0.0382** | 0.8002 | 0.3140 | **0.0145** | 0.5399 | 0.3467 | 0.1268 | 0.4656 | 0.3399 | 0.1779 | 0.4066 | 0.3386 | 0.2364 | 0.8032 | 0.2889 | **0.0080** |
| **Disease Severity (severe)** | 0.4684 | 0.2916 | 0.1155 | 0.2513 | 0.2924 | 0.3950 | 0.1253 | 0.3229 | 0.6999 | 0.4430 | 0.3166 | 0.1689 | 0.3634 | 0.3153 | 0.2554 | 0.3205 | 0.2691 | 0.2402 |
| **Days pso** | 0.0863 | 0.1410 | 0.5436 | 0.0462 | 0.1414 | 0.7453 | 0.1522 | 0.1561 | 0.3351 | 0.0701 | 0.1531 | 0.6491 | 0.0480 | 0.1524 | 0.7542 | 0.0118 | 0.1301 | 0.9282 |

Est (estimates), SE (standard error), pso (post symptom onset)

#### Table S2e. Generalised linear regression models of CD4 T cell cytokine responses in SARS-CoV-2 recovered patients

| | CD4 Spike IFN-γ | | | CD4 Spike IL-2 | | | CD4 Spike TNF | | | CD4 M+N IFN-γ | | | CD4 M+N IL-2 | | | CD4 M+N TNF | | |
|---|---|---|---|---|---|---|---|---|---|---|---|---|---|---|---|---|---|---|
| **Coefficient** | **Est** | **SE** | **p value** | **Est** | **SE** | **p value** | **Est** | **SE** | **p value** | **Est** | **SE** | **p value** | **Est** | **SE** | **p value** | **Est** | **SE** | **p value** |
| **Age** | 0.3170 | 0.2982 | 0.3128 | 0.2555 | 0.2459 | 0.3233 | 0.2092 | 0.3048 | 0.5081 | 0.2674 | 0.3133 | 0.4135 | 0.1944 | 0.2594 | 0.4708 | -0.3688 | 0.3110 | 0.2632 |
| **Sex (male)** | 0.3723 | 0.6867 | 0.5996 | 1.0306 | 0.5663 | 0.0988 | 0.3834 | 0.7020 | 0.5970 | 0.2759 | 0.7215 | 0.7102 | 0.5908 | 0.5973 | 0.3460 | -0.2361 | 0.7162 | 0.7485 |
| **DM (yes)** | -0.7169 | 0.5252 | 0.2022 | -0.4192 | 0.4331 | 0.3560 | -0.4518 | 0.5369 | 0.4198 | -0.7481 | 0.5518 | 0.2050 | -0.3923 | 0.4568 | 0.4106 | 0.0420 | 0.5478 | 0.9403 |
| **Obesity (yes)** | 0.1716 | 0.6298 | 0.7908 | 0.1389 | 0.5193 | 0.7945 | 0.8563 | 0.6438 | 0.2130 | 0.5275 | 0.6617 | 0.4438 | -0.0329 | 0.5478 | 0.9533 | -0.1901 | 0.6568 | 0.7782 |
| **Disease Severity (severe)** | 0.3686 | 0.5900 | 0.5462 | 0.1065 | 0.4865 | 0.8312 | -0.4126 | 0.6031 | 0.5094 | -0.1854 | 0.6199 | 0.7710 | 0.2750 | 0.5132 | 0.6037 | -0.1665 | 0.6154 | 0.7922 |
| **Days pso** | -0.2078 | 0.3253 | 0.5373 | -0.3230 | 0.2683 | 0.2562 | 0.0027 | 0.3325 | 0.9936 | -0.1130 | 0.3418 | 0.7478 | -0.5099 | 0.2830 | 0.1017 | -0.4725 | 0.3393 | 0.1939 |
| Est (estimates), SE (standard error), pso (post symptom onset) | | | | | | | | | | | | | | | | | | |

#### Table S2f. Generalised linear regression models of CD8 T cell cytokine responses in SARS-CoV-2 recovered patients

| | CD8 Spike IFN-γ | | | CD8 Spike IL-2 | | | CD8 Spike TNF | | | CD8 M+N IFN-γ | | | CD8 M+N IL-2 | | | CD8 M+N TNF | | |
|---|---|---|---|---|---|---|---|---|---|---|---|---|---|---|---|---|---|---|
| **Coefficient** | **Est** | **SE** | **p value** | **Est** | **SE** | **p value** | **Est** | **SE** | **p value** | **Est** | **SE** | **p value** | **Est** | **SE** | **p value** | **Est** | **SE** | **p value** |
| **Age** | -0.3468 | 0.3103 | 0.2899 | -0.5560 | 0.2141 | **0.0266** | -0.2412 | 0.3162 | 0.4633 | -0.0623 | 0.3305 | 0.8542 | -0.5717 | 0.2919 | 0.0786 | -0.2982 | 0.2994 | 0.3427 |
| **Sex (male)** | -0.4403 | 0.7146 | 0.5516 | 0.1160 | 0.4930 | 0.8187 | -0.6419 | 0.7282 | 0.3987 | -0.1148 | 0.7610 | 0.8831 | 0.0510 | 0.6722 | 0.9410 | 0.3871 | 0.6893 | 0.5868 |
| **DM (yes)** | -0.6703 | 0.5465 | 0.2481 | -0.2953 | 0.3771 | 0.4516 | 0.2779 | 0.5569 | 0.6286 | -0.0871 | 0.5820 | 0.8840 | -0.1144 | 0.5141 | 0.8283 | 0.0486 | 0.5272 | 0.9284 |
| **Obesity (yes)** | -0.0194 | 0.6553 | 0.9770 | 1.2171 | 0.4521 | **0.0226** | 0.6295 | 0.6678 | 0.3680 | 0.6008 | 0.6979 | 0.4095 | 0.3194 | 0.6164 | 0.6156 | 0.6140 | 0.6322 | 0.3543 |
| **Disease Severity (severe)** | 0.6468 | 0.6140 | 0.3169 | 0.6714 | 0.4236 | 0.1441 | 0.6328 | 0.6256 | 0.3356 | -0.7529 | 0.6538 | 0.2763 | 0.2345 | 0.5775 | 0.6933 | -0.3396 | 0.5923 | 0.5790 |
| **Days pso** | 0.0369 | 0.3385 | 0.9154 | 0.3537 | 0.2335 | 0.1608 | 0.2361 | 0.3449 | 0.5091 | -0.0864 | 0.3605 | 0.8155 | -0.2600 | 0.3184 | 0.4332 | 0.0121 | 0.3265 | 0.9711 |

Est (estimates), SE (standard error), pso (post symptom onset)

**Table S3. Antibodies used for intracellular cytokine staining and proliferation assay**

| Marker | Fluorochrome | Clone | Isotype | Species reactivity | Host Species | Manufacturer | Catalogue number | Dilution used |
|---|---|---|---|---|---|---|---|---|
| CD14 | APC-Fire750 | M5E2 | IgG2a, κ | Human | Mouse | Biolegend, UK | 301854 | 200 |
| CD3 | PerCP | UCHT1 | IgG1, κ | Human | Mouse | Biolegend, UK | 300428 | 100 |
| CD3 | FITC | UCHT1 | IgG1, κ | Human | Mouse | Biolegend, UK | 300440 | 50 |
| CD4 | APC | RPA-T4 | IgG1, κ | Human | Mouse | Biolegend, UK | 300514 | 200 |
| CD8 | PE-Cy7 | RPA-T8 | IgG1, κ | Human | Mouse | Biolegend, UK | 301012 | 200 |
| CD8 | BV510 | RPA-T8 | IgG1, κ | Human | Mouse | Biolegend, UK | 301048 | 600 |
| IFN-γ | PE | 4S.B3 | IgG1, κ | Human | Mouse | Biolegend, UK | 502508 | 50 |
| IL-2 | PE-Cy7 | MQ1-17H12 | IgG2a, κ | Human | Rat | eBioscience, USA | 25-7029-41 | 100 |
| TNF | FITC | MAb11 | IgG1, κ | Human | Mouse | Biolegend, UK | 502906 | 40 |

### Methods

***Generalised linear models***

Generalised linear models (GLMs) were performed to estimate the association of obesity (categorical) and DM (categorical) with antibody, B cell, and T cell immune responses (all continuous and log-transformed) while adjusted for age (continuous), sex (discrete), disease severity (categorical) and days post symptoms onset (continuous) in SARS-CoV-2 recovered patients. All continuous variables were standardised by z-score normalisation. Normality of the data was tested by Shapiro–Wilk test, histogram, and Q-Q diagnostic plots. Interactions and co-linearity between variables were explored.

Model < - glm (immune response ∼ age + sex + diabetes status + obesity category + disease severity + days post symptoms onset, data = data)
